# A novel scale for assessing caregiving competence in family caregivers of persons with dementia

**DOI:** 10.1101/2024.03.10.24304060

**Authors:** Ippei Suganuma, Noriyuki Ogawa, Kenji Kamijou, Aki Nakanishi, Ippei Kawasaki, Keisuke Itotani, Shinichi Okada

**Author notes:** Corresponding author (IS).

## Abstract

The aging of family caregivers and the challenges of long-distance caregiving attributed to the increase in the number of elderly individuals living alone have raised concerns about dementia caregiving in Japan. Additionally, with the shifts in family dynamics due to declining birth rates and an extended average lifespan, adapting support strategies for family caregivers is necessary. Thus, it is necessary to measure the caregiving competence of family caregivers early and effectively. However, a comprehensive caregiving competence scale tailored to dementia, including aspects such as caregiving burden, affirmation, and coping, is lacking. Therefore, this study aimed to develop a Caregiving Competence Scale for Dementia (CCSD) for primary family caregivers caring for individuals with dementia. This study focused on primary family caregivers caring for individuals with cognitive impairment and various degrees of dementia. The initial version of the CCSD was developed, and a questionnaire survey was conducted to validate its structural validity and reliability. A total of 150 participants were included in the analysis. The exploratory factor analysis identified five factors with 27 items: Factor 1: “Positive Emotions and Awareness,” Factor 2: “Presence or Absence of Consultation Partners and Family Support,” Factor 3: “Caregiving Burden and Coping Skills,” Factor 4: “Dementia Literacy,” and Factor 5: “Engagement and Emotional Control.” The confirmatory factor analysis revealed a good model fit (comparative fit index = 0.905 and root mean square error of approximation = 0.072). The overall Cronbach’s alpha coefficient for the scale was 0.892. The CCSD, comprising 27 items covering five factors, has been successfully developed as a measurement scale. Measuring caregiving competence contributes to developing targeted support strategies for primary family caregivers and facilitating appropriate interventions.

## Introduction

According to the World Health Organization, over 55 million people worldwide have dementia, with approximately 10 million new cases reported each year [1]. Dementia has profound physical, psychological, social, and economic impacts, affecting not only individuals with dementia but also their caregivers, families, and society as a whole [1]. In Japan, with a population of approximately 120 million, the elderly population rate is 29.1% (about 36 million) [2], and the prevalence of dementia is estimated to be 20% (about 7 million) by 2025 [3]. Moreover, dementia is the leading cause of needing caregiving assistance (18.0%) among individuals aged 65 years and older [4]. With the aging population in Japan, the number of caregivers for persons with dementia is expected to increase.

The burden on dementia caregivers has been reported to be significantly higher than that on other caregivers, leading to lower levels of self-efficacy, subjective well-being, and physical health [5]. This burden is influenced by various factors, such as behavioral and psychological symptoms of dementia (BPSD) [6–9], physical dependence in activities of daily living [10], and background factors, including the caregiver–patient relationship [6], living arrangements [6,11], caregiver gender [12,13], kinship [11], and social support [13].

Recently, in the context of family caregivers in Japan, issues related to aging caregivers and long-distance caregiving attributed to the increase in the number of elderly individuals living alone have been emphasized [14]. Additionally, changes in family dynamics due to the aging population and extended average lifespan have altered household situations and the characteristics of primary family caregivers [14]. Thus, primary family caregivers must flexibly address challenges that arise in their daily lives while striving to coordinate and construct a home caregiving environment.

Considering the challenges family caregivers face in Japan, developing a measurement scale for the caregiving competence of family caregivers of persons with dementia and mild cognitive impairment (MCI) is necessary for providing early and appropriate support to family caregivers. Although there is no consistent definition for caregiving competence, in caregiving practice, negative emotions, such as caregiver burden, coexist with positive emotions, such as self-efficacy [15], facilitating appropriate caregiving responses to the care recipient [16,17]. Therefore, in developing the scale for caregiving competence, it is crucial to define the core concept of “caregiving competence” by encompassing both positive and negative emotions and coping strategies in caregiving situations.

Although no scales have been developed for measuring caregiving competence, an Empowerment Evaluation Scale for caregivers has been developed as a comprehensive measure of caregiving abilities [18,19]. The scale developed by Wu [18] includes various aspects, such as caregiving autonomy, consciousness, and relationships with care recipients, but it is not specialized for dementia caregiving. The scale developed by Sakanashi et al. [19] is specific to family caregivers of persons with dementia. However, this scale does not include stress, coping, and family relationships, giving the impression of inadequacy in capturing the multifaceted nature of caregiving competence. Therefore, this study aimed to develop a new scale for measuring caregiving competence in primary family caregivers of persons with dementia or MCI by reexamining the elements that constitute “caregiving competence.”

## Materials and methods

### Operational definition of caregiving competence

In this study, the operational definition of caregiving competence was “the ability to provide continuous care for persons with dementia or MCI at home by adjusting to the environment (family, local residents, use of caregiving services, etc.), addressing BPSD, and providing physical care related to activities of daily living, including supervision.”

### Development of the CCSD

Based on the operational definition, items from previous studies on caregiving-related assessment scales [18,19], caregiving survey reports [20], and interviews with family caregivers were considered. Responses to questions were rated on a 5-point scale, with 5 indicating “Strongly agree (always or frequently),” 4 indicating “Somewhat agree (often or somewhat),” 3 indicating “Neither agree nor disagree (Neither always nor rarely or Neither frequently nor rarely),” 2 indicating “Somewhat disagree (rarely or not much),” 1 indicating “Strongly disagree (never).” The higher the scores, the higher the care competence (reverse scoring for negative emotions or situations). As a result, 45 items were generated, covering caregiving burden, caregiving affirmation, self-efficacy for knowledge, availability of a confidant, resources, coping skills, and balancing caregiving with work. The generated items were reviewed for face validity with 15 family primary caregivers of persons with MCI or dementia. After face validity confirmation, the content validity was reviewed by five experts (university faculty and medical professionals specializing in dementia care and welfare), resulting in the creation of the “Caregiving Competence Scale for Dementia Prototype (CCSD-P)” (Table 1).

**Table 1.**
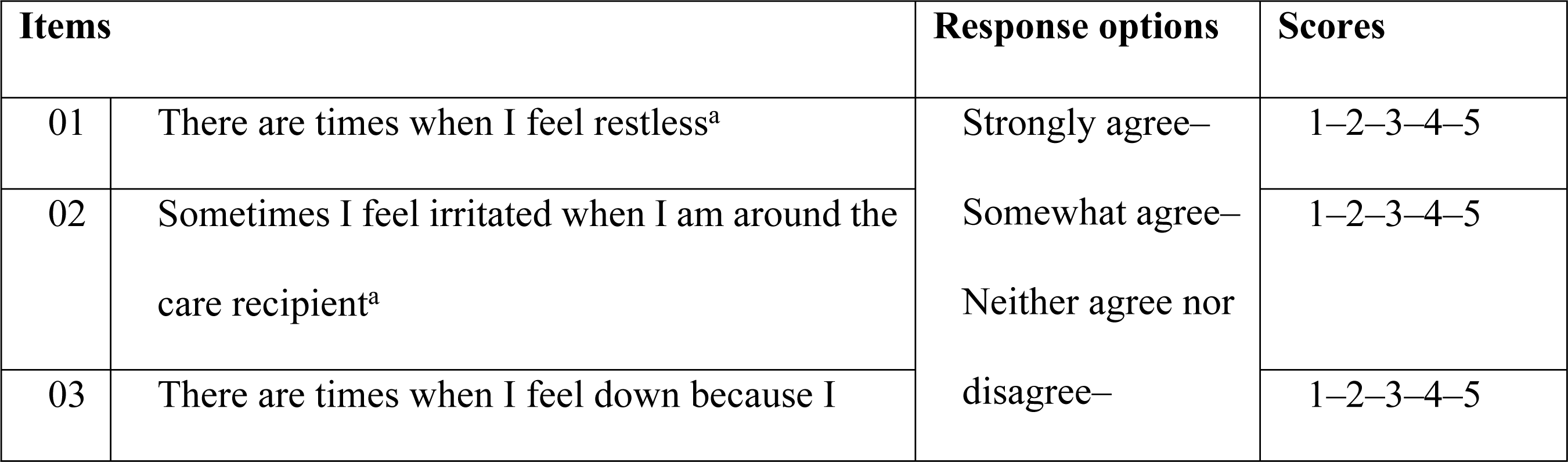
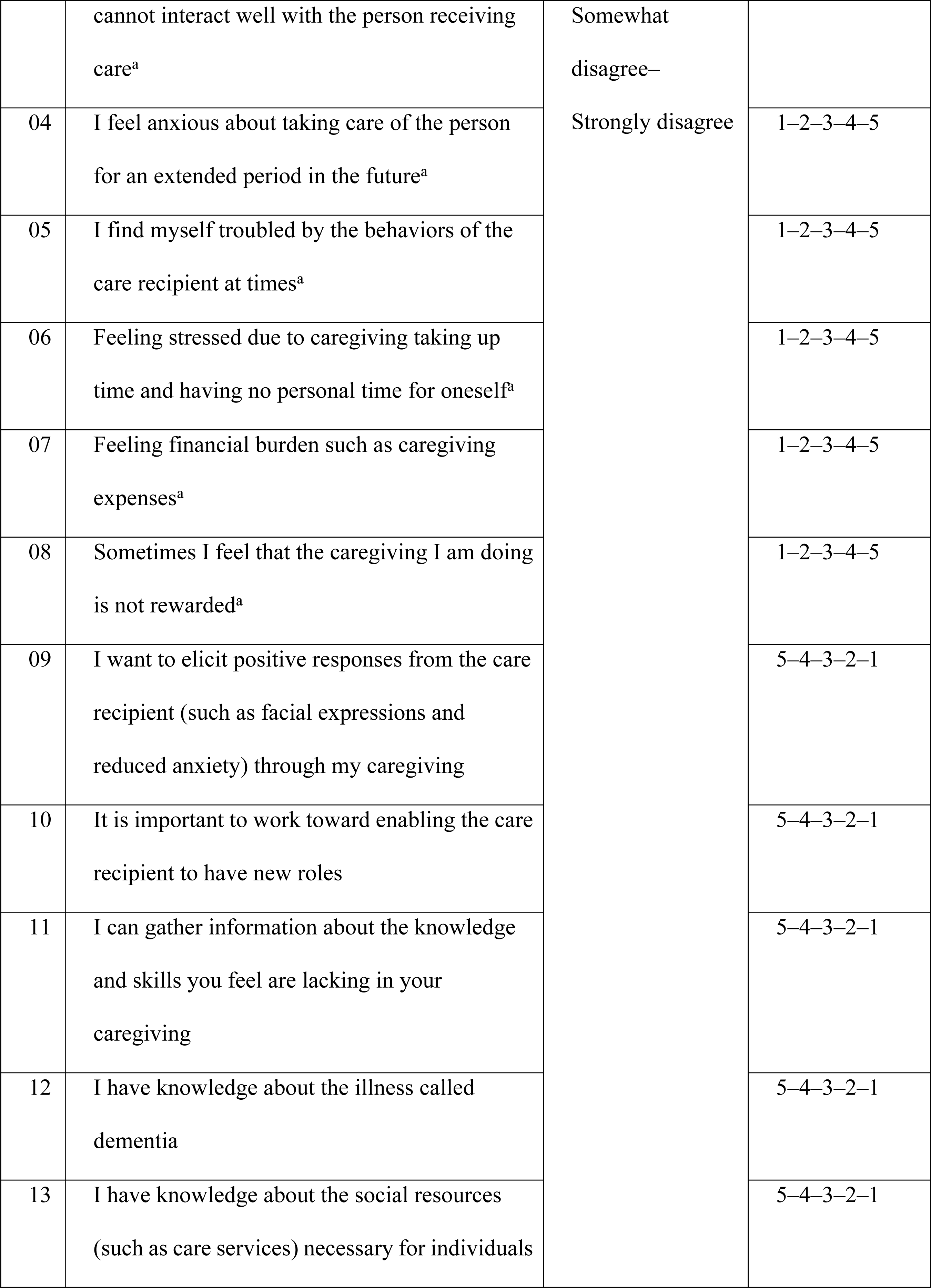
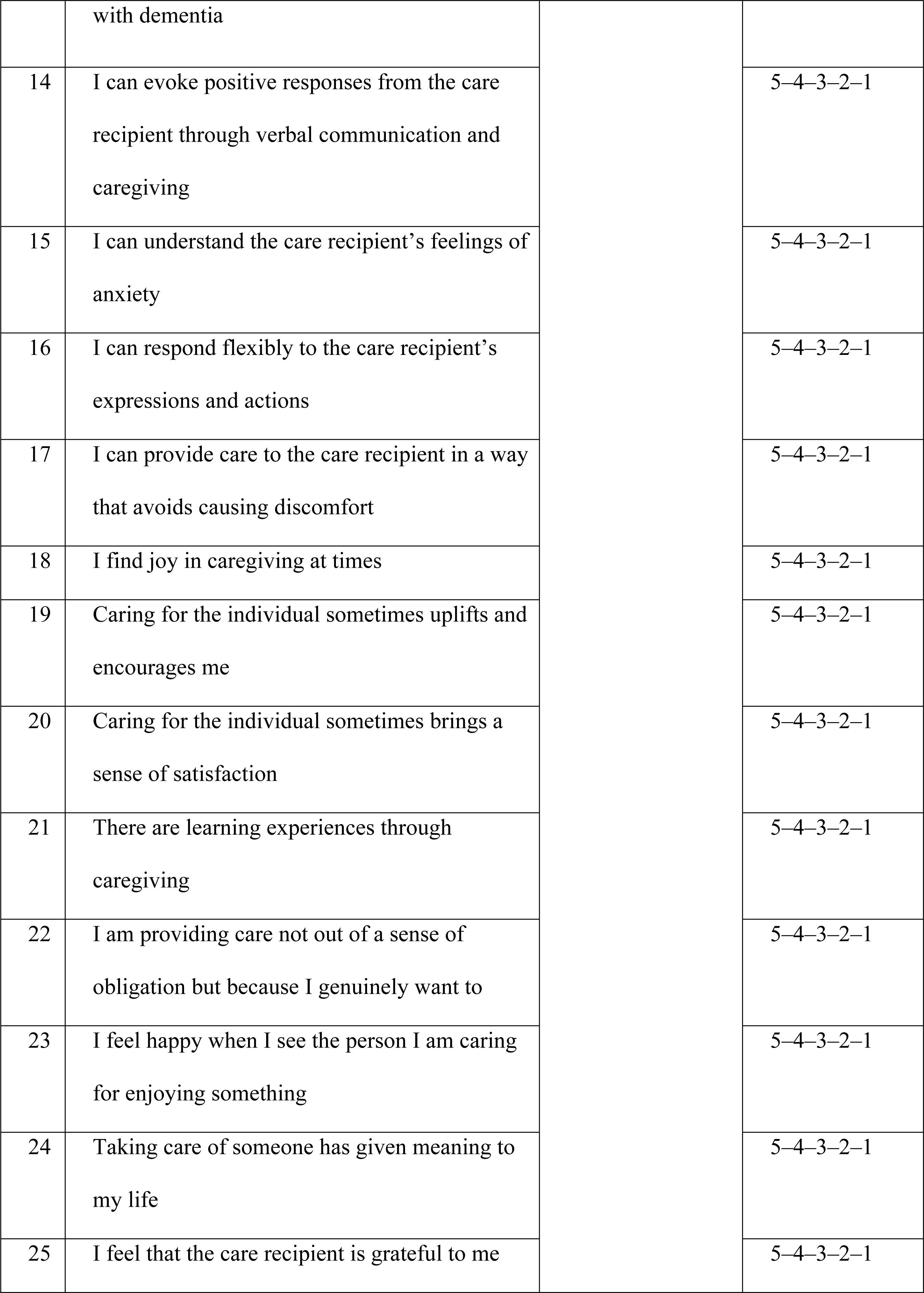
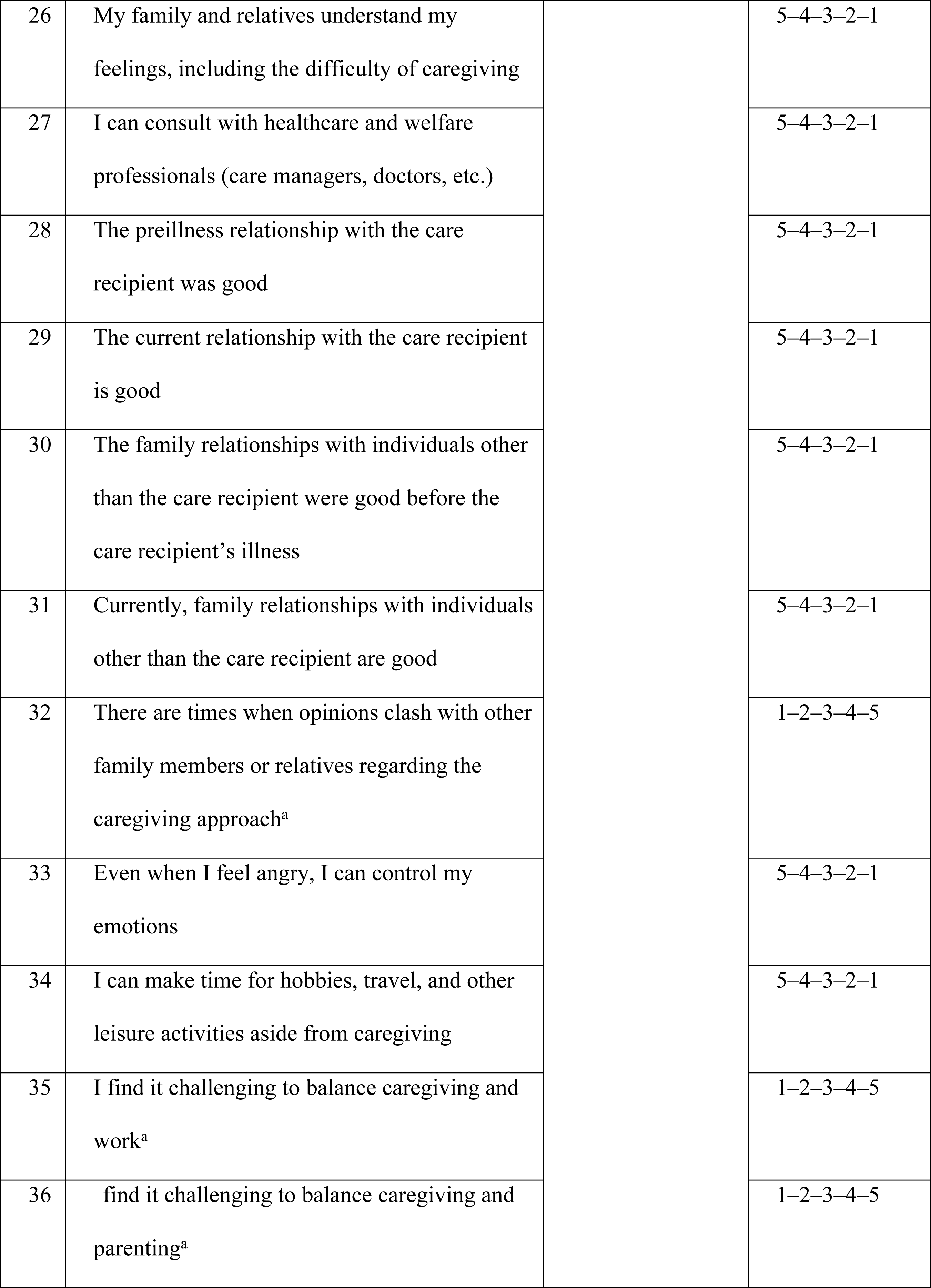
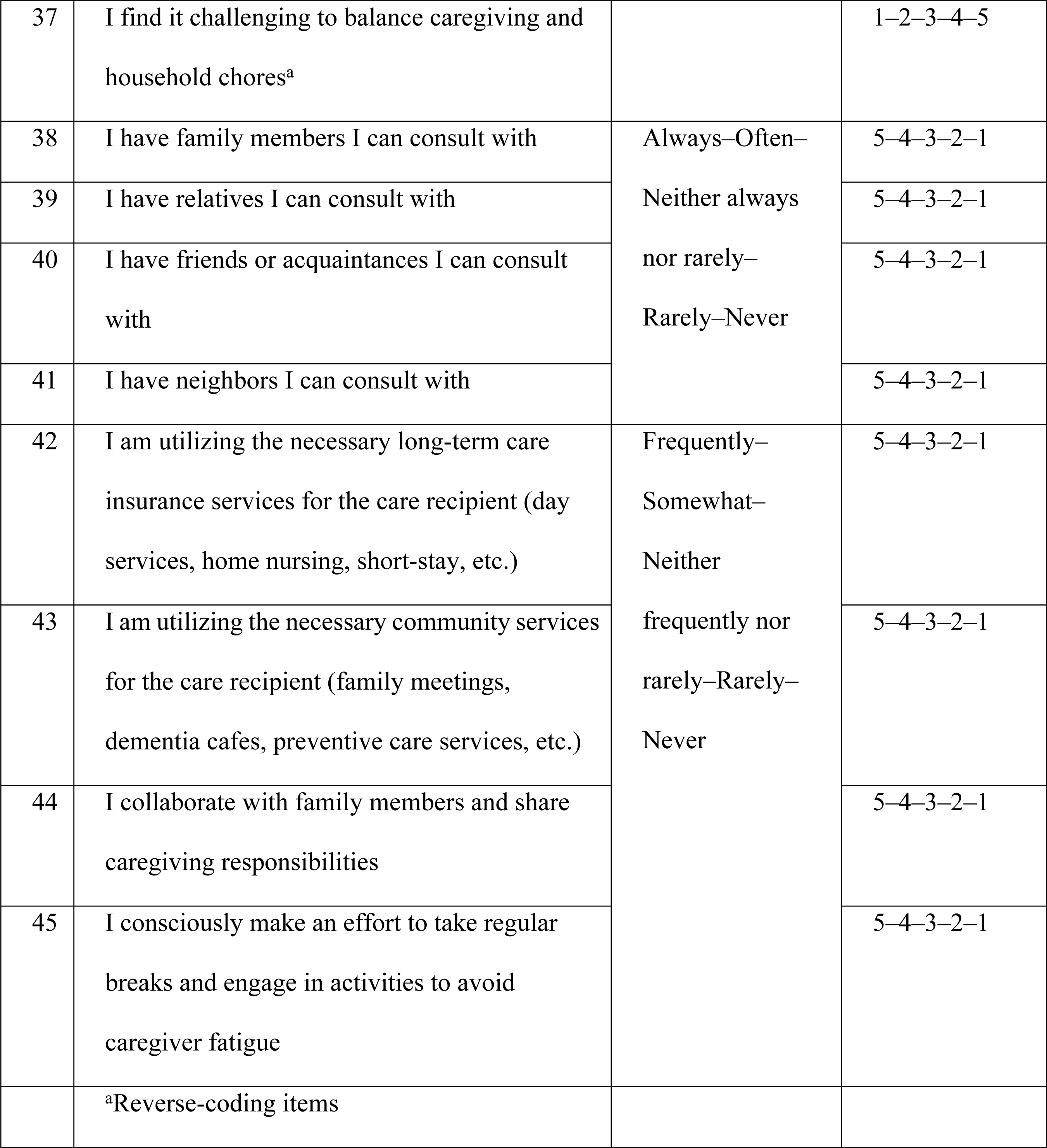
Caregiving Competence Scale for Dementia Prototype (CCSD-P)

### Participants

The survey targeted caregivers providing care for persons with MCI or mild-to-severe dementia for at least 25 min per day. The diagnosis of dementia was considered, even in cases where the specific type of dementia was unclear, but the caregiver was informed by a medical professional about the presence of dementia symptoms. The caregiving time criterion was based on the care required according to the Japanese Ministry of Health, Labour and Welfare standards. Caregiving included both direct physical care and supervision in daily life. Both cohabiting and noncohabiting caregivers were included.

### Instruments

A questionnaire was administered to collect demographic information about caregivers (age, gender, relationship with the care recipient, and education duration). Previous studies have reported associations between education duration and caregiver grief [21] and burden [22]. Thus, the education duration was included. Education duration was categorized as “9 years or less,” “10–12 years” (up to high school graduation or equivalent), and “13 years or more” (post-high school education). Additionally, in dementia caregiving, depressive symptoms are frequently observed in caregivers [23]. Therefore, the Japanese version of the Center for Epidemiologic Studies Depression Scale (CES-D) [24] was included as a survey item to assess depressive symptoms. The CES-D is a self-rating scale for depressive symptoms over the past week, comprising 20 items rated on a 4-point scale with a total score of 60. A cutoff score of 16 or higher indicates stronger depressive symptoms.

Information about care recipients was collected, including age, gender, diagnosis, and frequency of BPSD assessed using the Japanese version of the Dementia Behavior Disturbance Scale short form (DBD-13) [25]. The DBD-13 consists of 13 items, each assessing the frequency of BPSD over the past month on a 5-point scale. The total score is 52.

Additional caregiving situation information was collected, including the number of cohabitants with the care recipient, daily caregiving time, duration since the diagnosis of dementia, duration of caregiving, number of caregiving service utilizations, and presence of a secondary caregiver. These survey items, along with the CCSD-P, were compiled into a questionnaire.

### Procedures

The survey was conducted in collaboration with the “Alzheimer’s Association Japan” (AAJ), which has branches in all 47 prefectures. A total of 23 branches were randomly selected, and the questionnaire was sent by mail to each branch. The distribution of the questionnaire to each branch was determined based on the actual situation, ranging from 5 to 20 questionnaires. A selection criteria letter was enclosed, and the distribution of questionnaires was determined by the branch executives, who then distributed them to the eligible participants. A return envelope was also included, and collection was performed via postal mail.

### Survey period

The survey period, approved by the ethics committee, was from March 23, 2023, to the questionnaire collection end date on December 31, 2023.

### Data analysis

Ceiling and floor effects were checked to detect any response bias and to verify the validity and reliability of the returned draft of the Caregiving Response Scale. Items demonstrating ceiling or floor effects were subsequently eliminated.

Exploratory factor analysis was performed to elucidate the factor structure and finalize the items for each factor. Items with communalities of less than 0.4 were eliminated during exploratory factor analysis. After confirming that no items needed to be removed based on communalities, promax rotation (maximum likelihood method) was performed. Items with factor loading values below 0.4 were eliminated, and this process was iteratively repeated until no removal targets remained. The interpretability of the factor items was carefully examined to confirm the final factor structure. Additionally, items with factor loading values of 0.35 or higher for a nonprimary factor were considered indicative of ambiguity and marked for removal.

Confirmatory factor analysis was subsequently performed to assess the validity of the factor structure obtained through exploratory factor analysis. Fit indices, namely, the comparative fit index (CFI) and the root mean square error of approximation (RMSEA), were used as model fit indicators, with CFI of >0.90 indicating good fit, RMSEA of <0.05 indicating excellent fit, RMSEA of <0.08 indicating good fit, and RMSEA of <0.10 indicating acceptable fit [26].

For reliability assessment, Cronbach’s α reliability coefficients were calculated for the overall scale and each subscale to ensure internal consistency. Items with reliability coefficients below 0.7 in the subscales were considered for potential removal [27].

### Ethical considerations

This study was approved by the Kyoto Tachibana University Research Ethics Committee (Approval Number: 22-60, Approval Date: March 23, 2023). Verbal and written consent were obtained from the participating branches of the AAJ before data collection. Participants were informed about the study purpose, personal information protection policy, research participation details (emphasizing voluntariness and the absence of disadvantages for nonparticipation), and other relevant information both orally and in writing. Consent was considered granted upon the return of the questionnaire.

## Results

A total of 259 questionnaires were distributed to the AAJ branches, of which 156 (recovery rate 60.2%) were collected. Of the 156 questionnaires collected, 6 were excluded due to missing values or exclusion criteria. Finally, 150 participants were considered for the analysis (effective recovery rate 57.9%).

### Basic information

#### Caregiver characteristics

The average age of caregivers was 68.4 ± 9.5 years. Of the 150 caregivers, 55 were males (36.6%), and 95 were females (63.3%). The caregiver relationship with the care recipient included husband (n = 47, 31.3%), wife (n = 45, 30.0%), son (n = 8, 5.3%), daughter (n = 35, 23.3%), son-in-law (n = 1, 0.7%), daughters-in-law (n = 8, 5.3%), brother (n = 1, 0.7%), sister (n = 3, 2.0%), and others (n = 2, 1.3%). Education duration was ≤9 years for 6 individuals (4%), 10–12 years for 54 (36.0%), and ≥13 years for 90 (60.0%). The average CES-D score was 20.6 ± 10.9 points, with 54 individuals (36.0%) scoring <16 and 96 (64.0%) scoring ≥16 (Table 2).

**Table 2.**
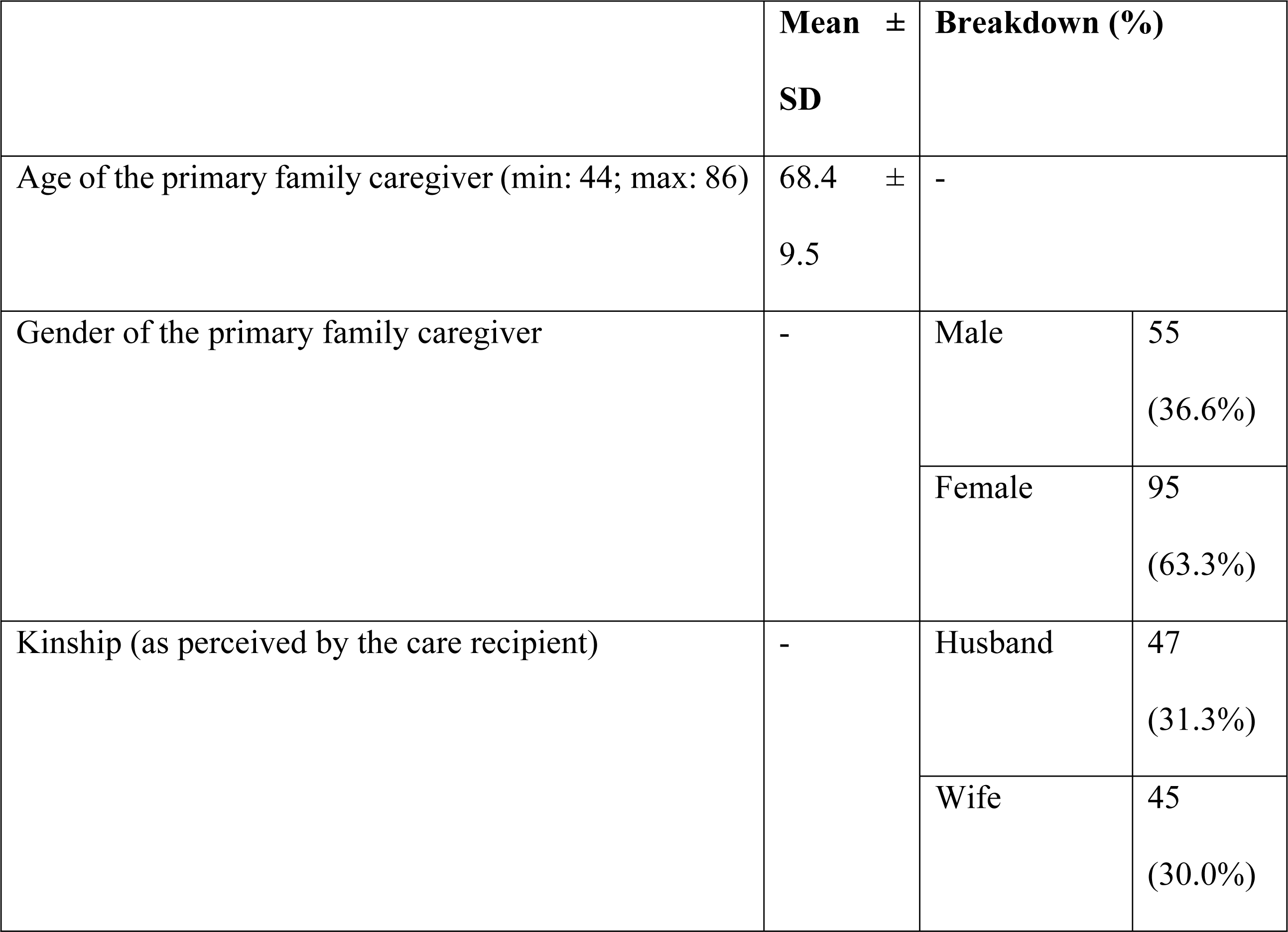
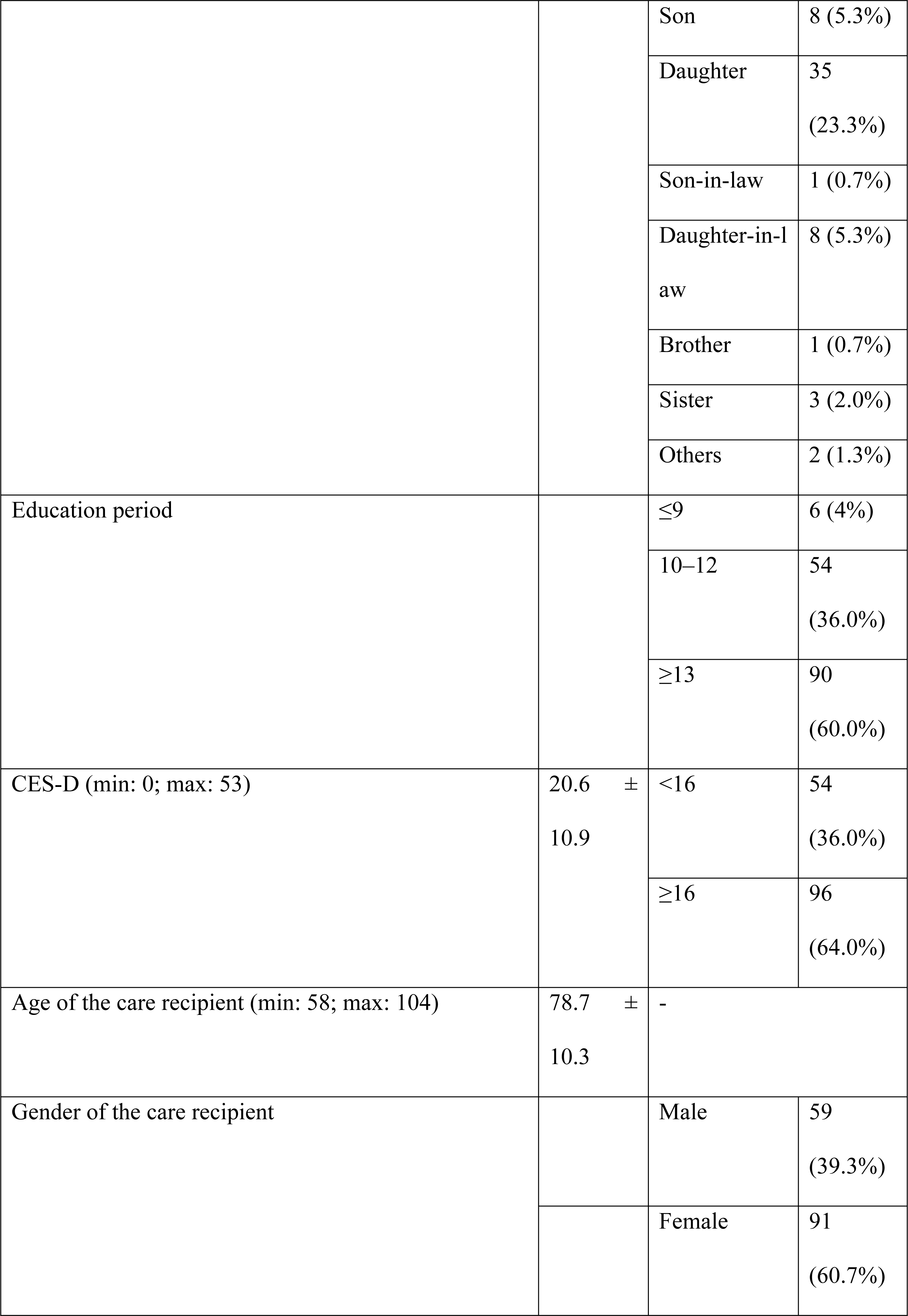
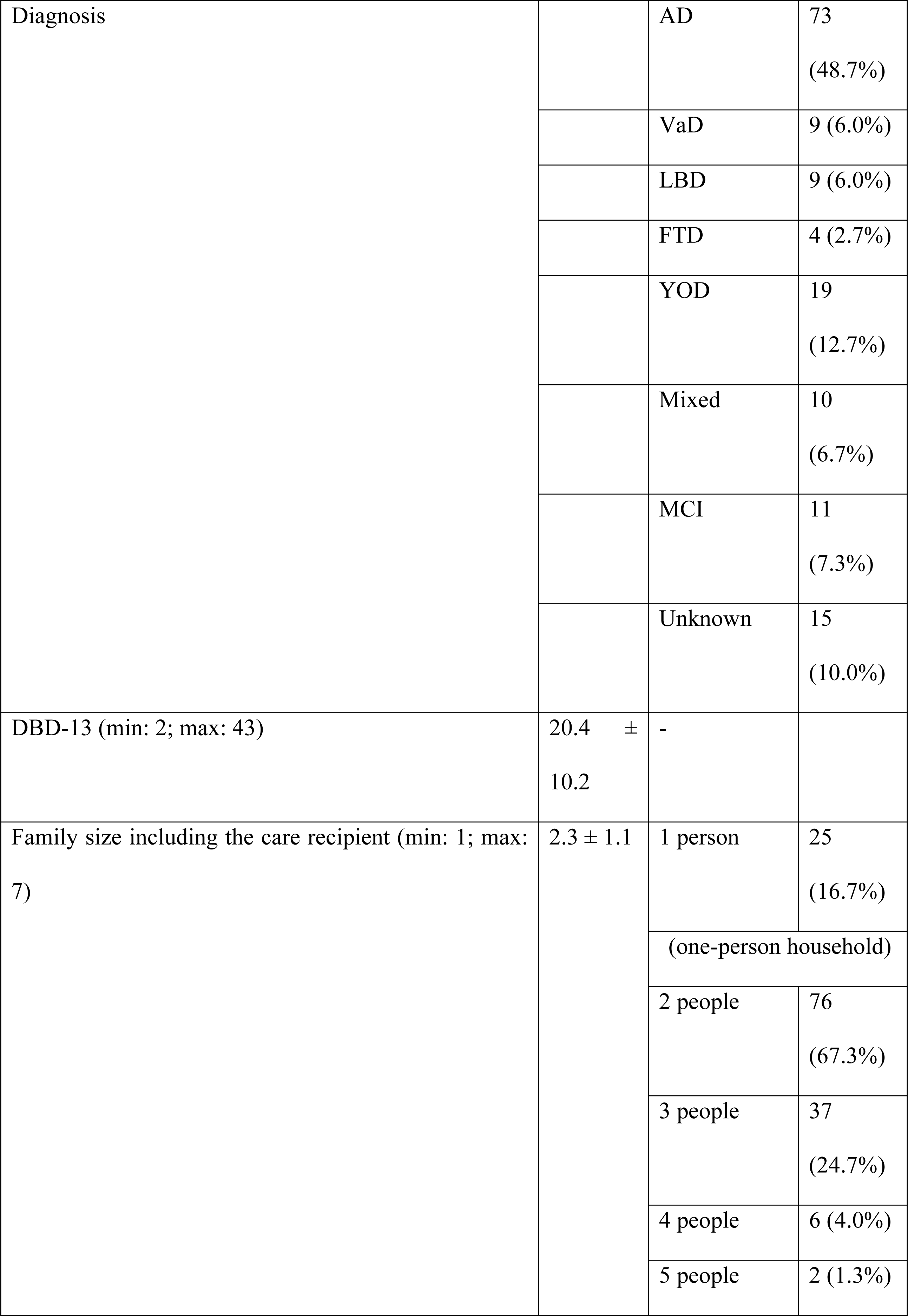
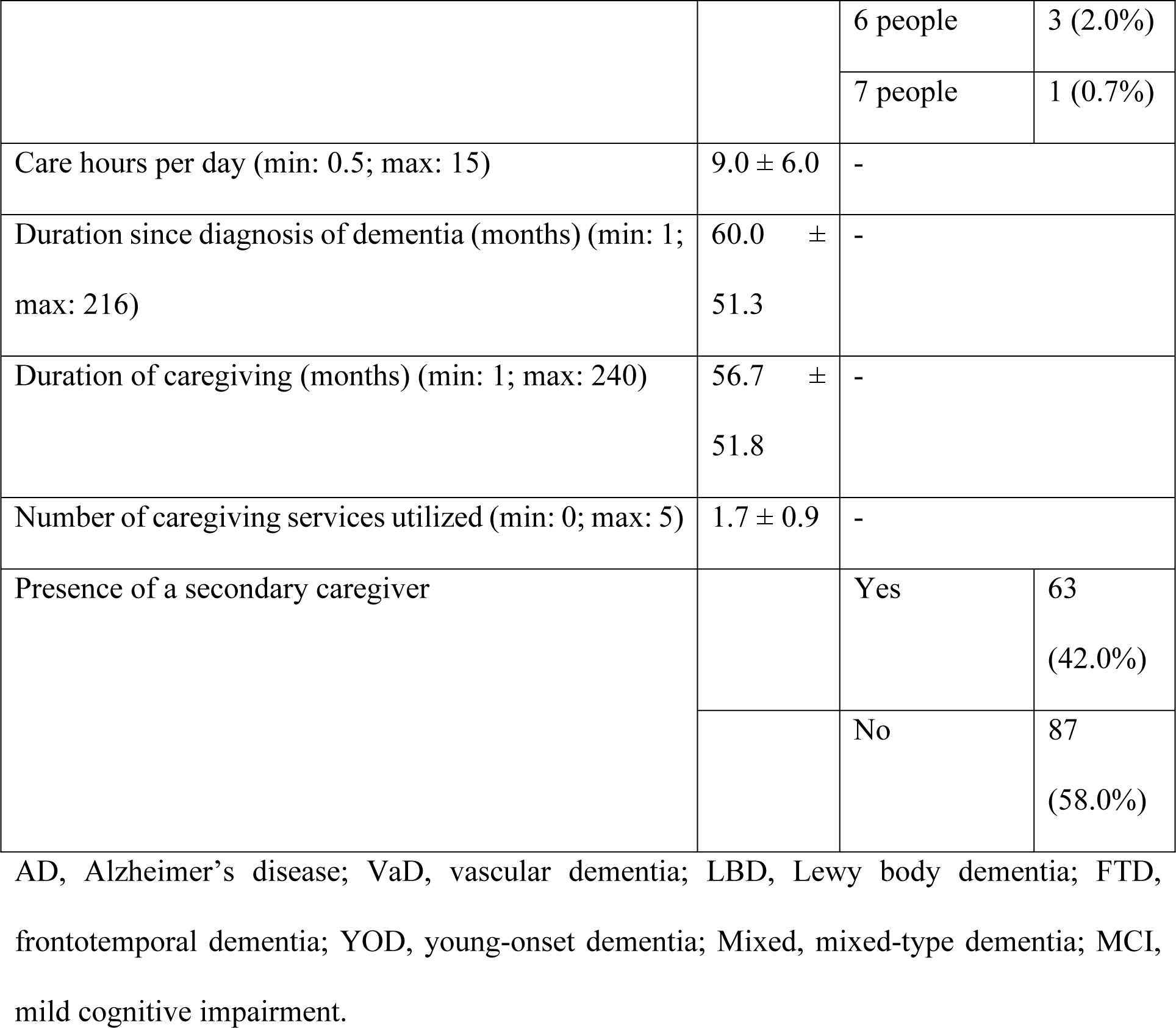
Basic information and attributes.

#### Care recipient characteristics

The average age of care recipients was 78.7 ± 10.3 years. Among them, 59 were males (39.3%), and 91 were females (60.7%). Of the 150 care recipients, 73 (48.7%) were diagnosed with Alzheimer’s disease (AD), 9 (6.0%) with vascular dementia, 9 (6.0%) with Lewy body dementia, 4 (2.7%) with frontotemporal dementia, 19 (12.7%) with young-onset dementia, 10 (6.7%) with mixed-type dementia, 11 (7.3%) with MCI, and 15 (10.0%) with unspecified type (diagnosed as dementia by a doctor without a specific diagnosis). The average DBD-13 score was 20.4 ± 10.2 points (Table 2).

#### Care situation

The average number of cohabitants for care recipients was 2.3 ± 1.1, and 25 (16.7%) care recipients were living alone. The average daily caregiving time was 9.0 ± 6.0 h, the average time since dementia diagnosis was 60.0 ± 51.3 months, the average caregiving duration was 56.7 ± 51.8 months, and the average number of services utilized was 1.7 ± 0.9. 42.0% of the care recipients (n = 63) had secondary caregivers, whereas 58.0% (n = 87) did not.

### Question items and ceiling/floor effects

Table 2 shows the scores and ceiling/floor effects of the 45 items considered in the CCSD-P. Items 4 and 5 showed floor effects, and item 42 showed a ceiling effect. Therefore, these items were eliminated (Table 3).

**Table 3.**
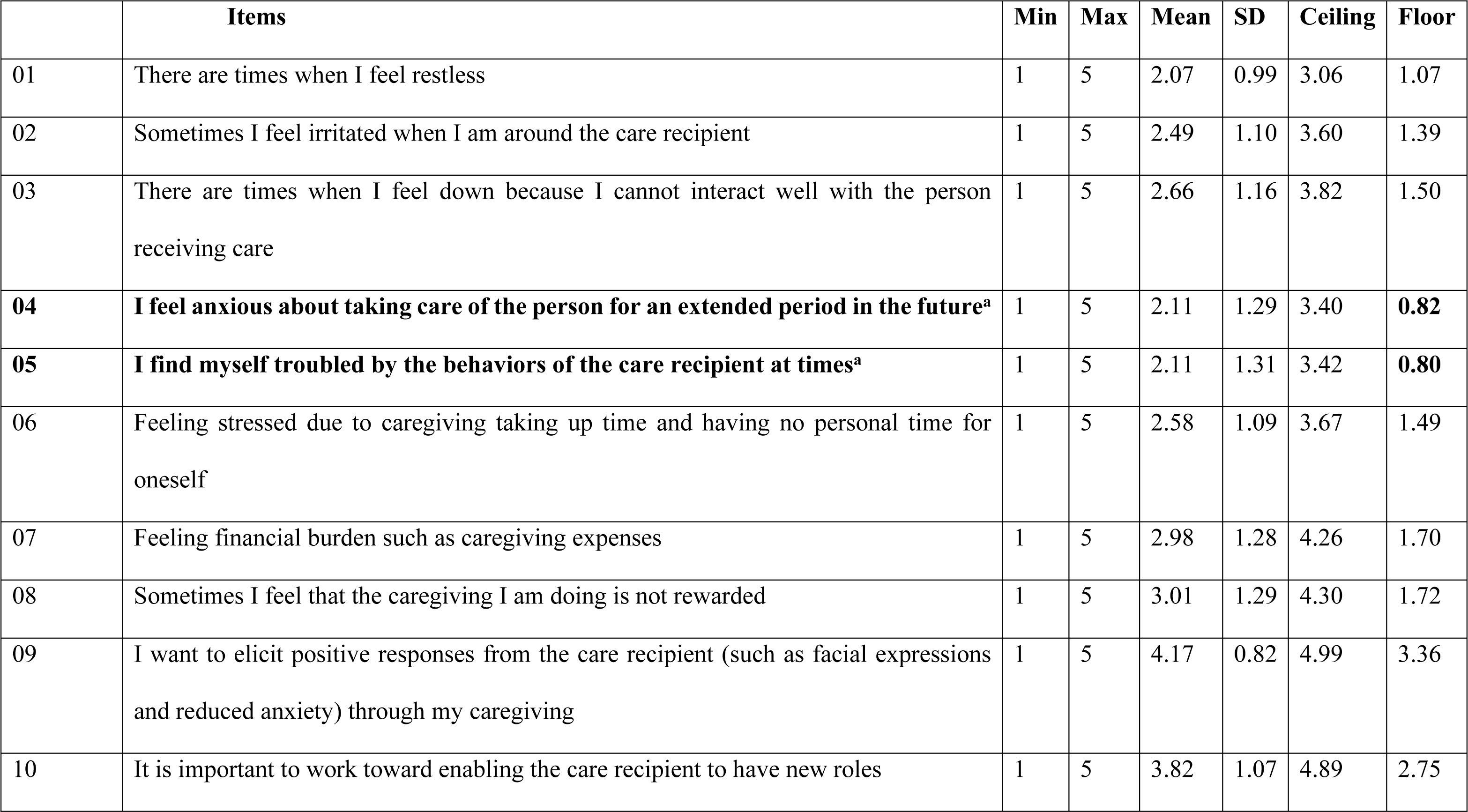
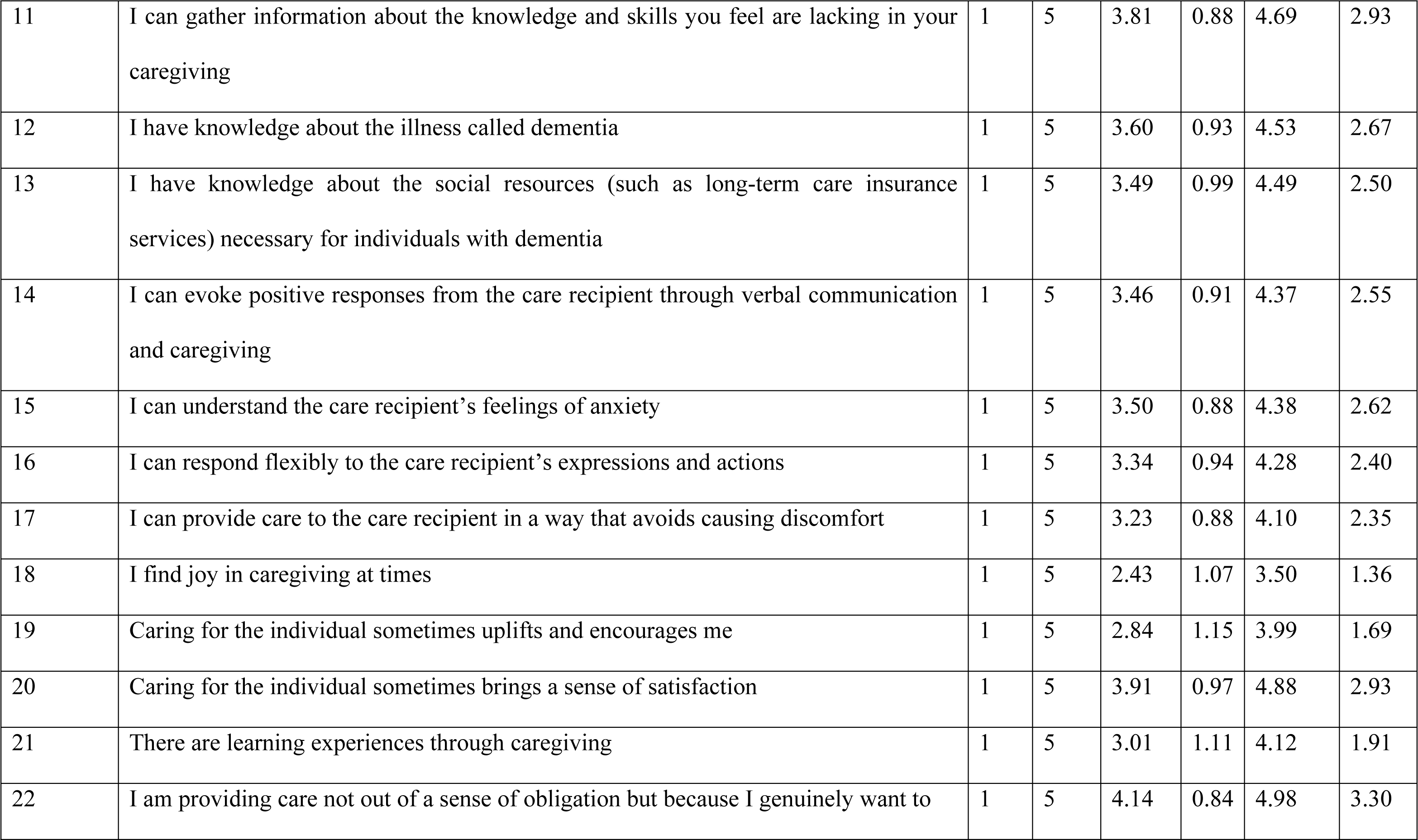
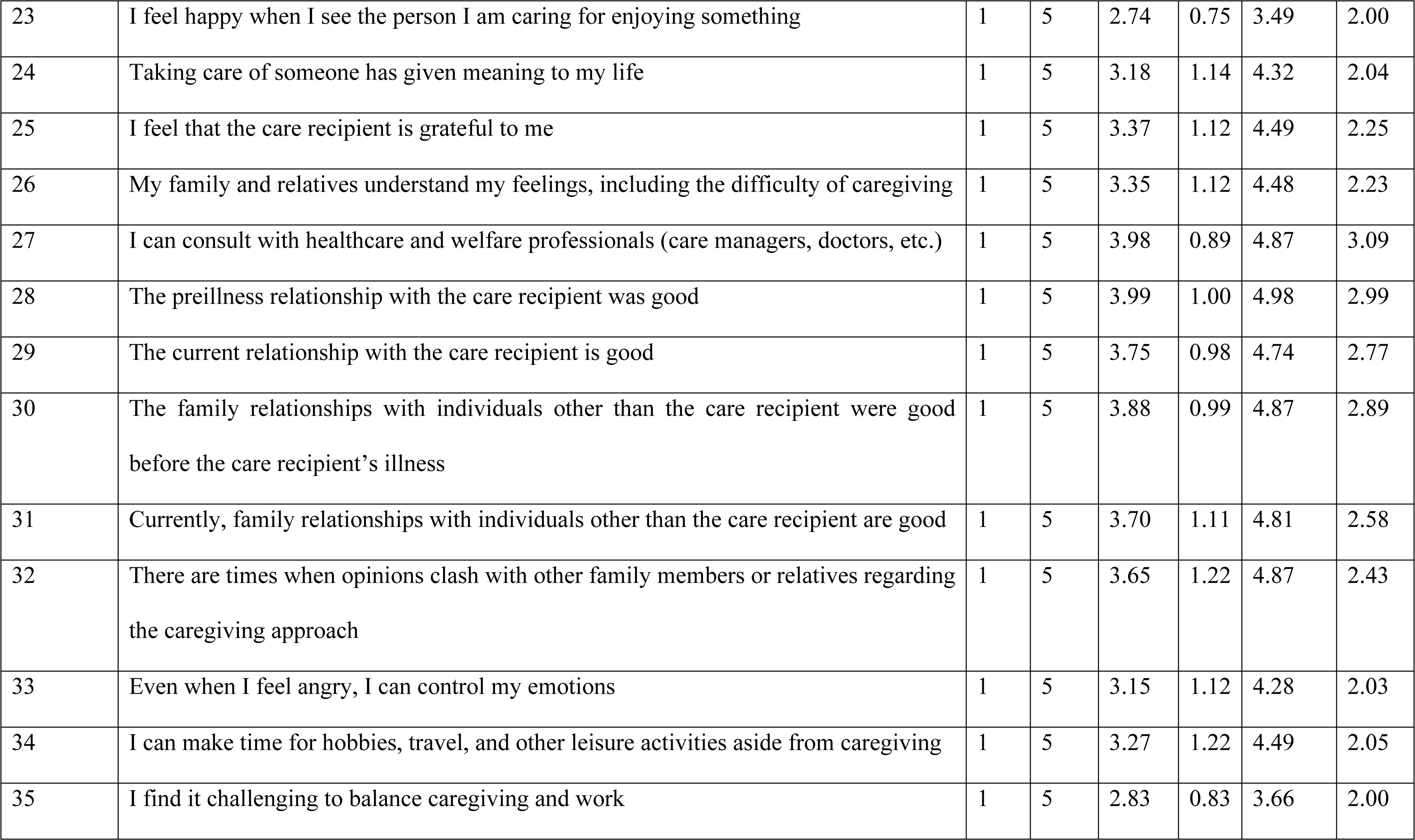
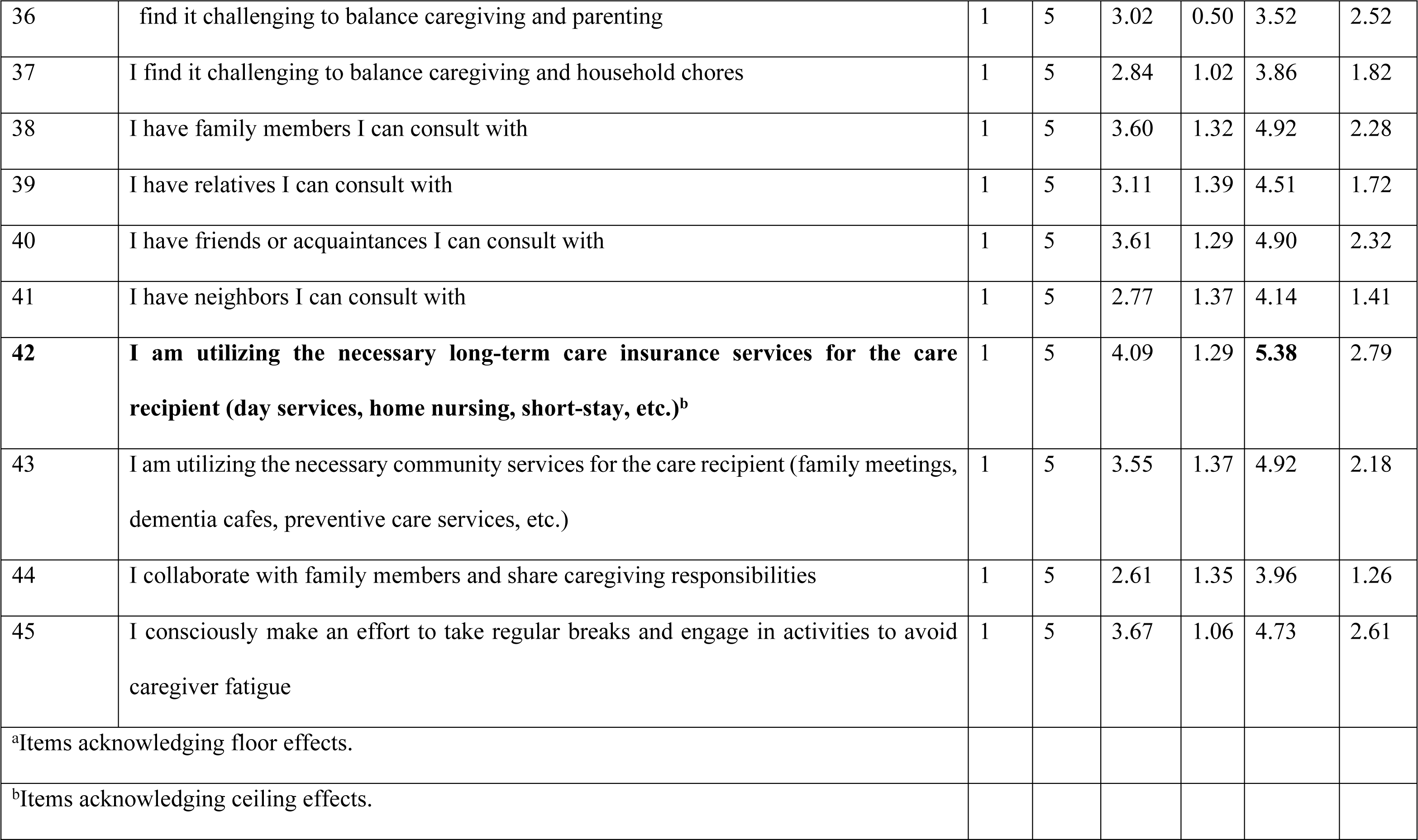
Mean scores and ceiling/floor effects of the CCSD-P questionnaire items.

### Results of exploratory and confirmatory factor analyses

Items 7, 9, 10, 21, 32, 35, 36, 37, 43, and 44 showed a commonality of less than 0.4 and were eliminated. Regarding the results of the scree plot, a five- or six-factor structure was considered. Based on the criterion of factor loading 0.40 or higher, items with low loading and those with ambiguity were eliminated. After repeated exploratory factor analysis, 27 items in the 5 factors were extracted. The Kaiser–Meyer–Olkin value was 0.851, and Bartlett’s sphericity test was significant at the 0.1% level, indicating that the scale has a statistically valid structure. The five-factor structure extracted by exploratory factor analysis was named “Positive Emotions and Awareness” for Factor 1, “Presence of Consultation Partners and Family Support” for Factor 2, “Care Burden and Coping Skills” for Factor 3, “Dementia Literacy” for Factor 4, and “Involvement and Emotion Control” for Factor 5 (Table 4 and Fig 1).

**Fig 1.**
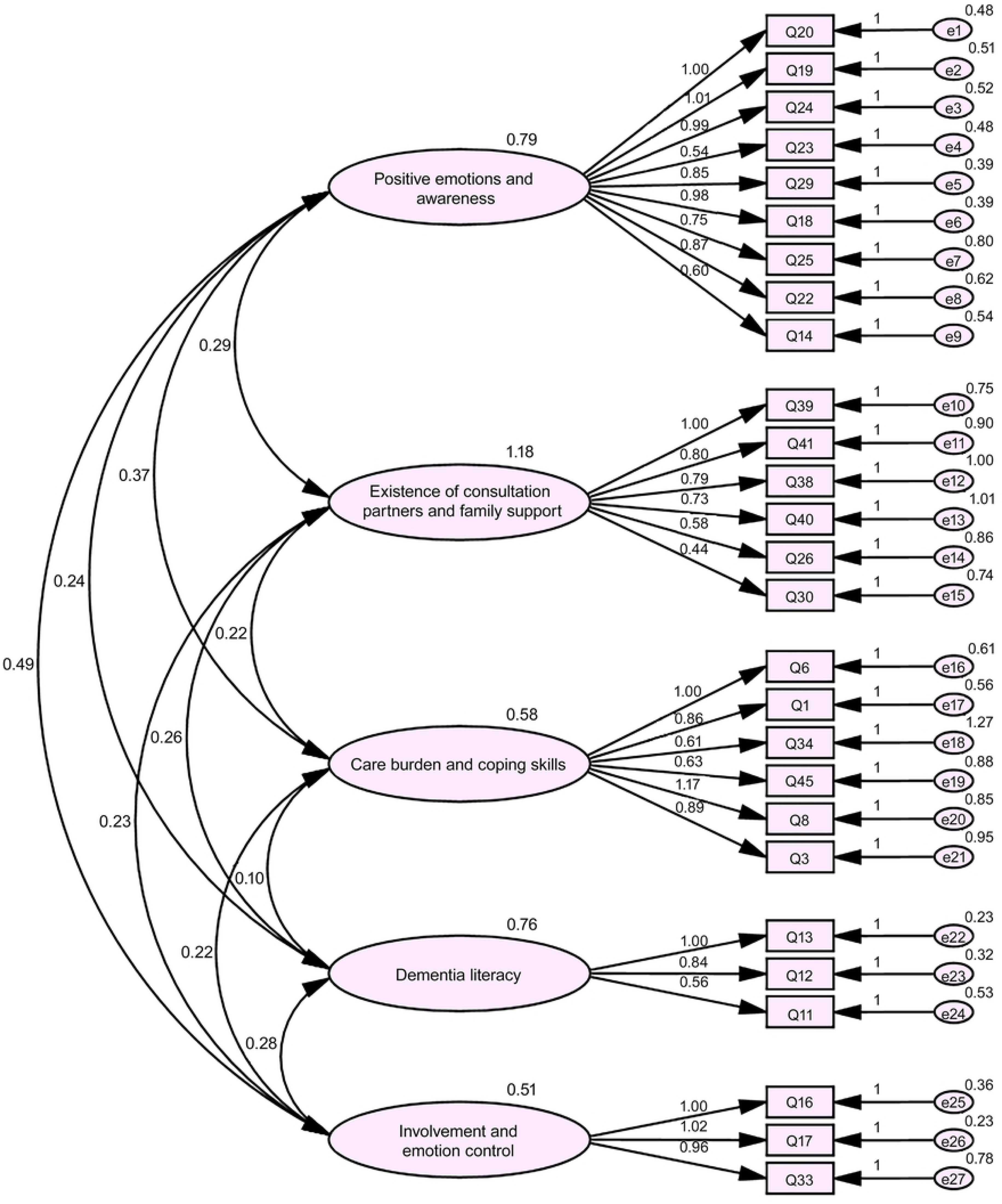
Results of confirmatory factor analysis. CFI = 0.905; RMSEA = 0.072.

**Table 4.**
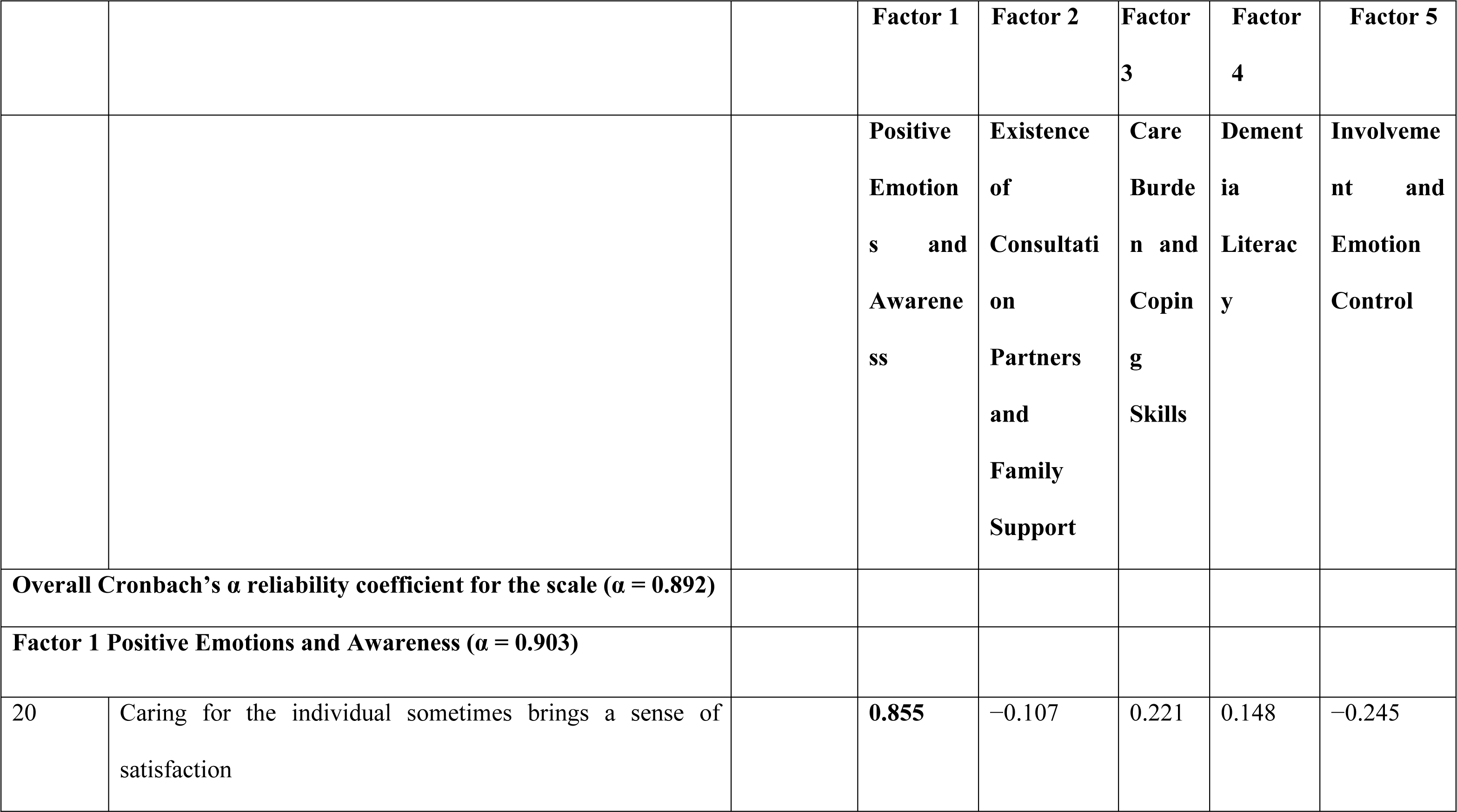
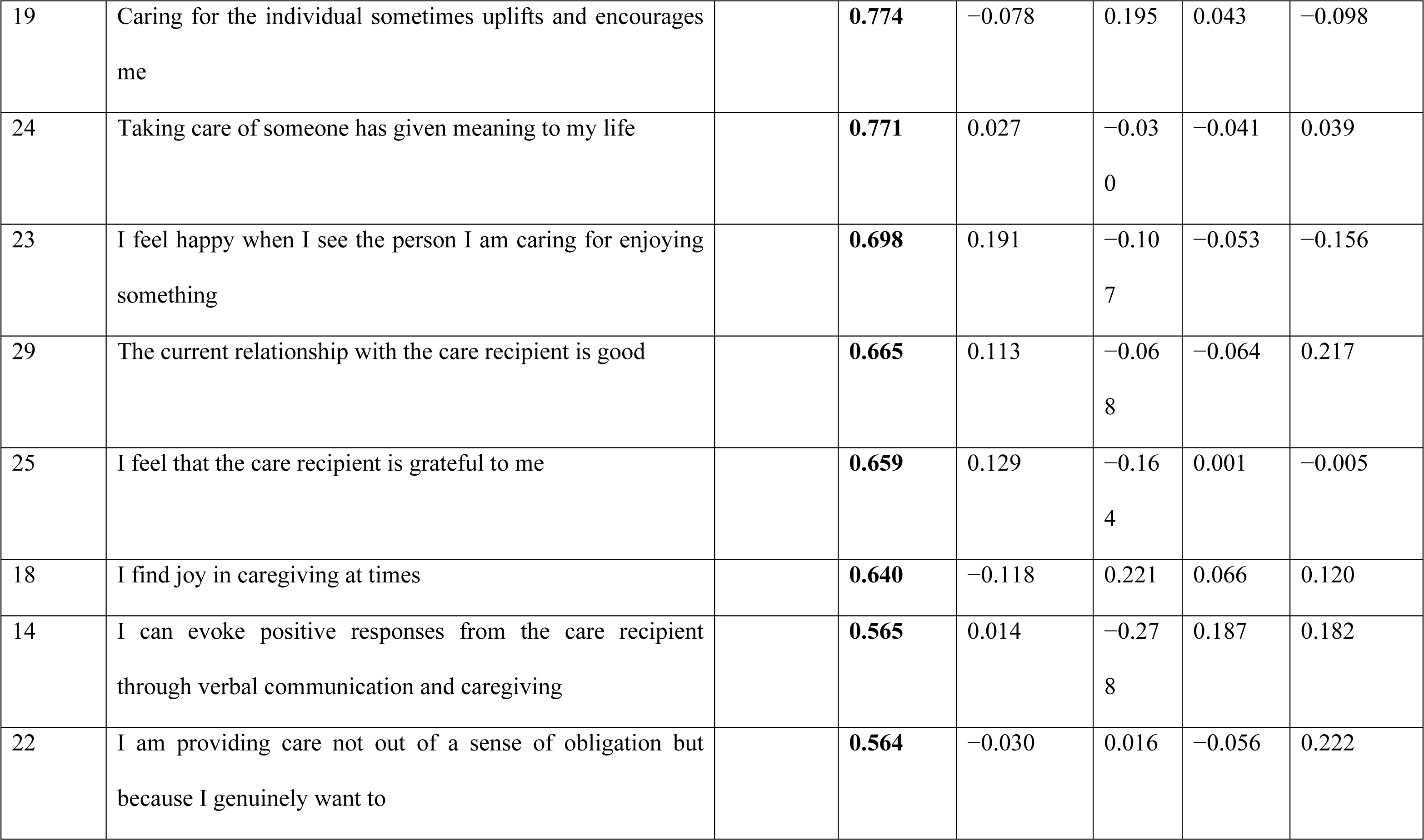
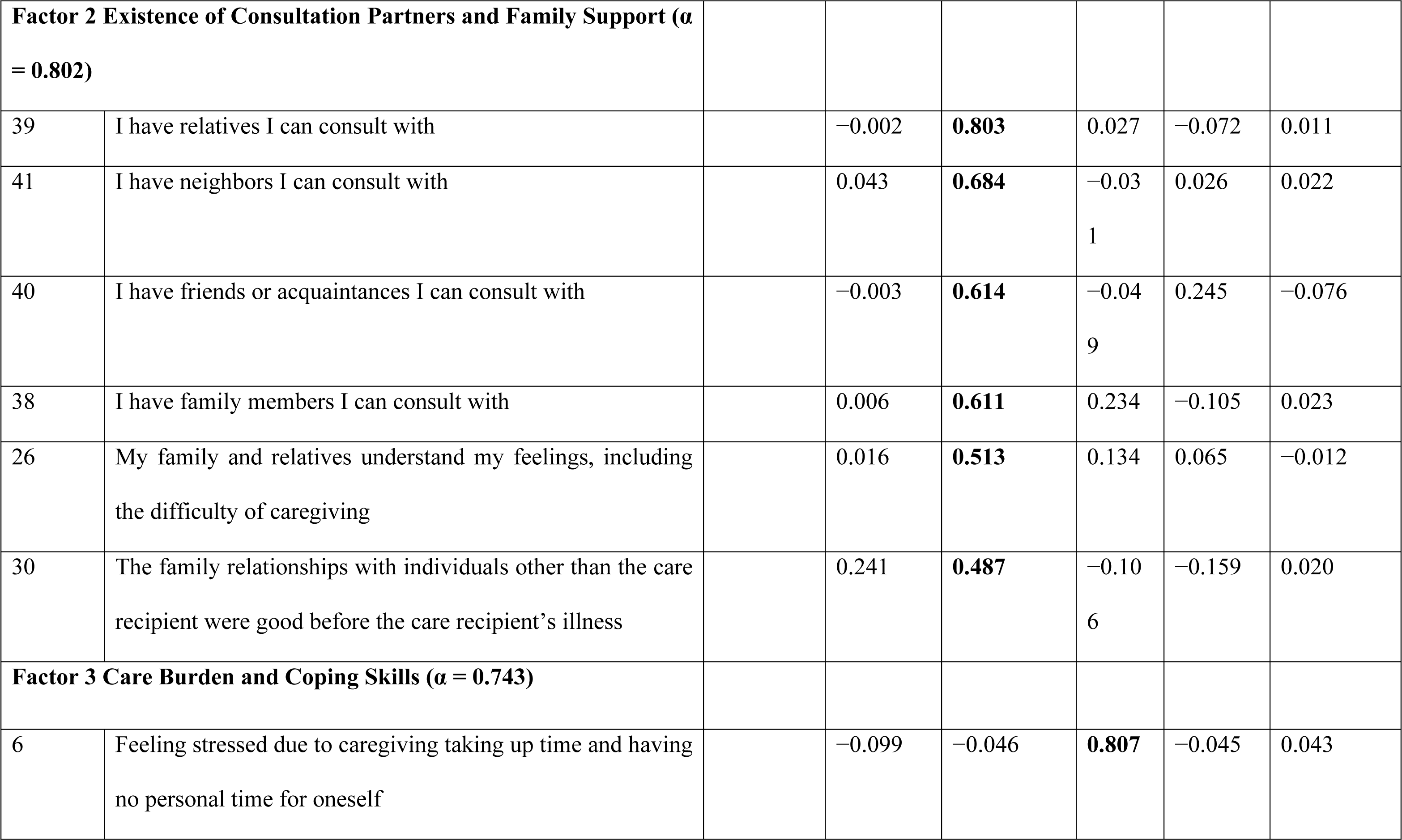
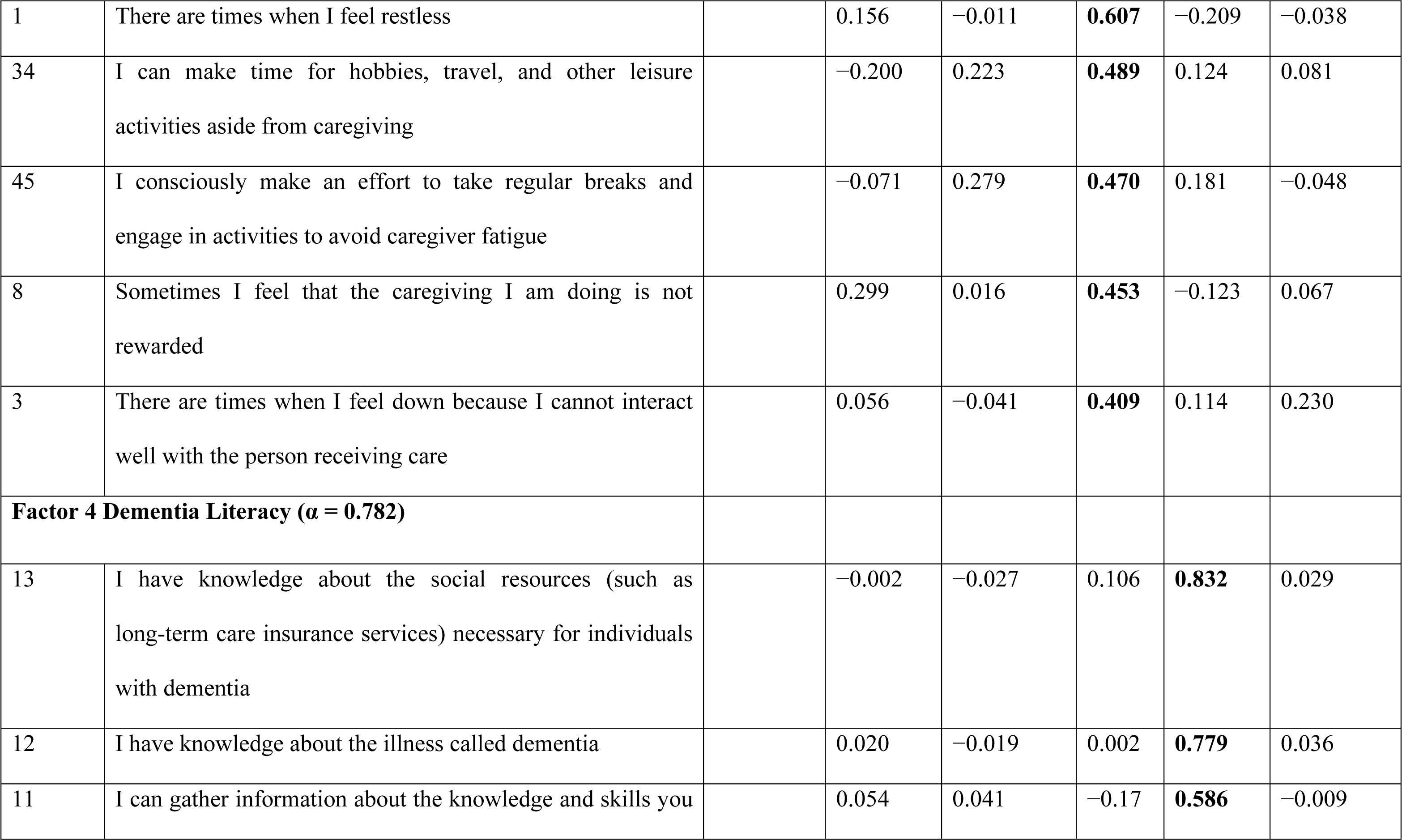
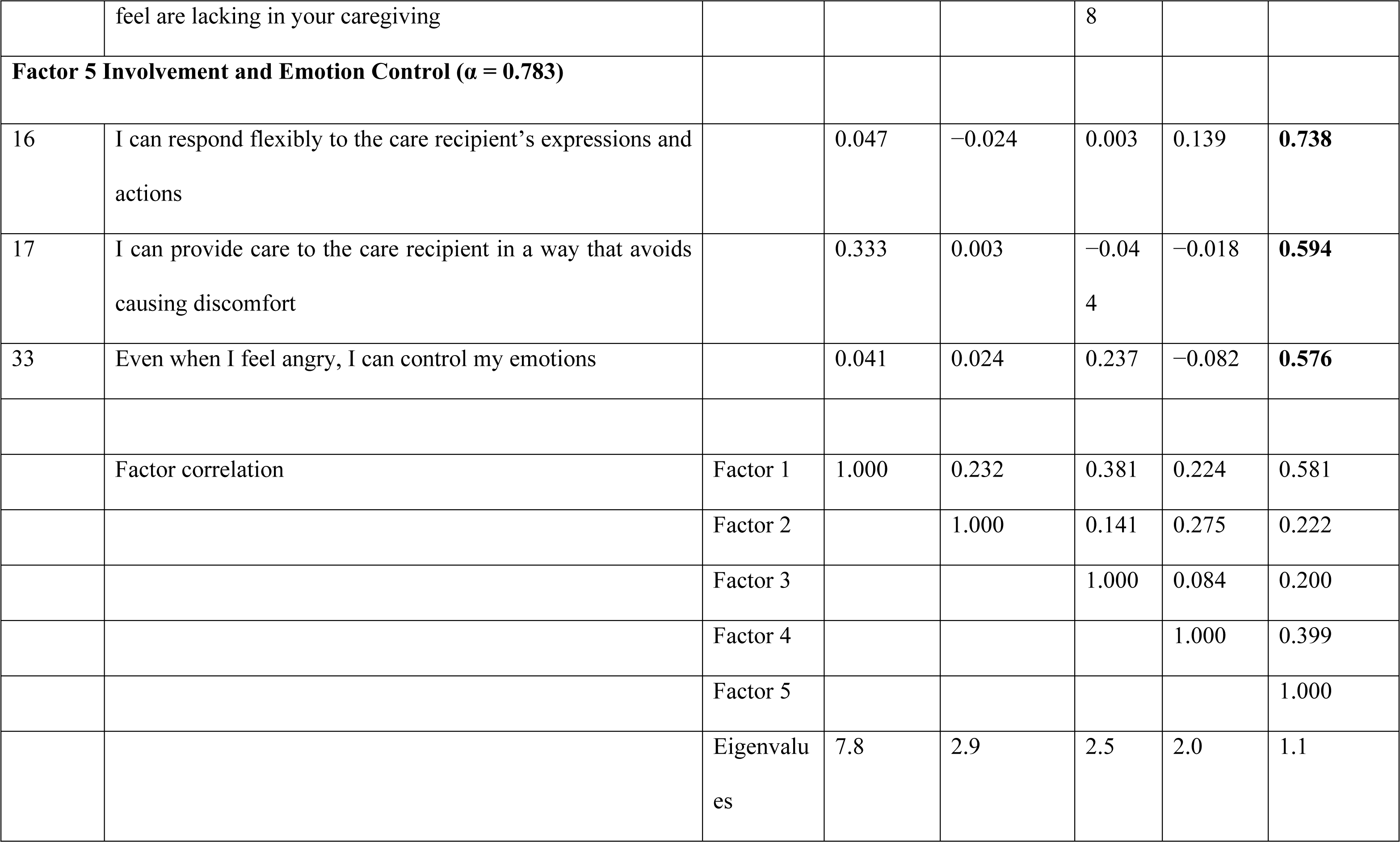
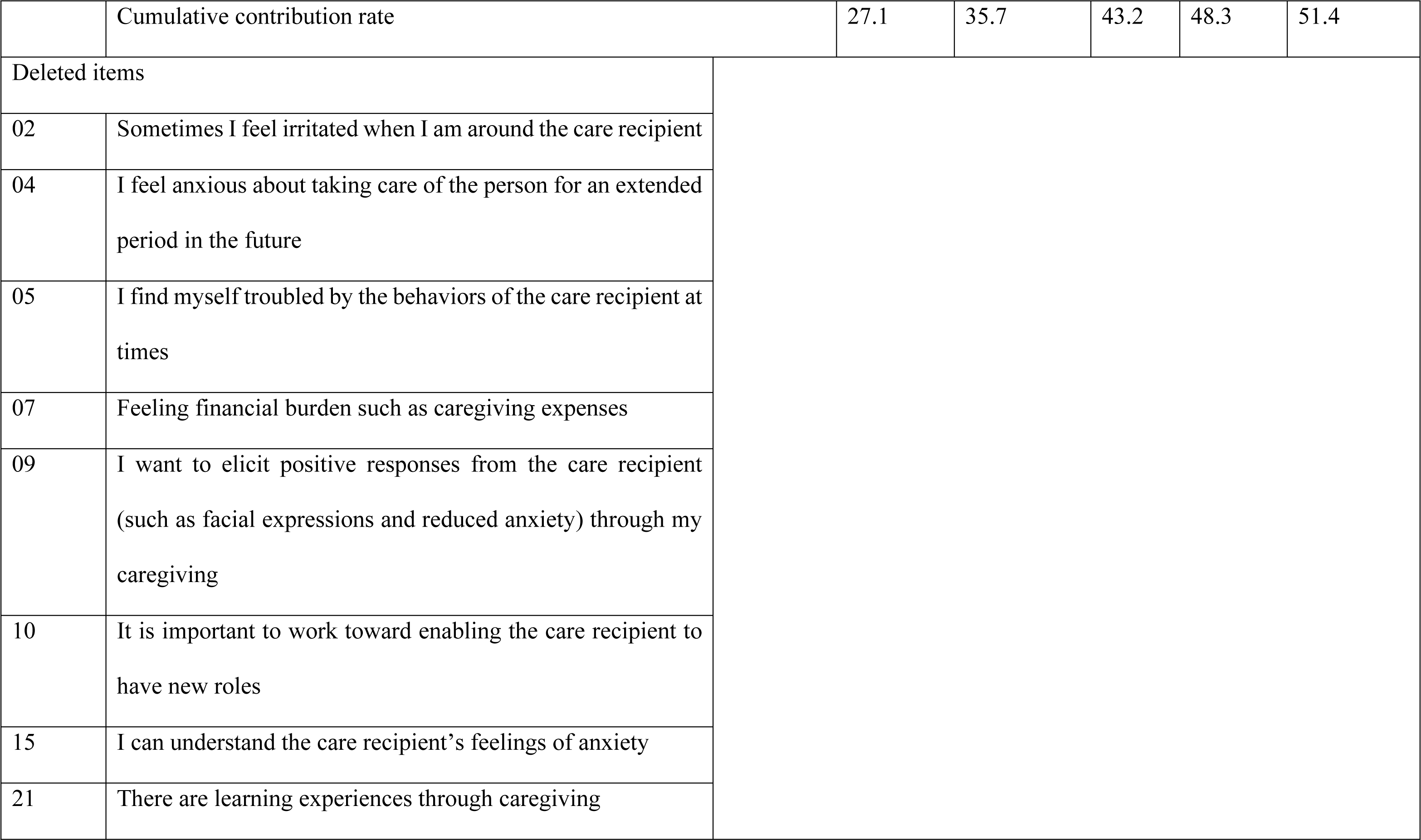
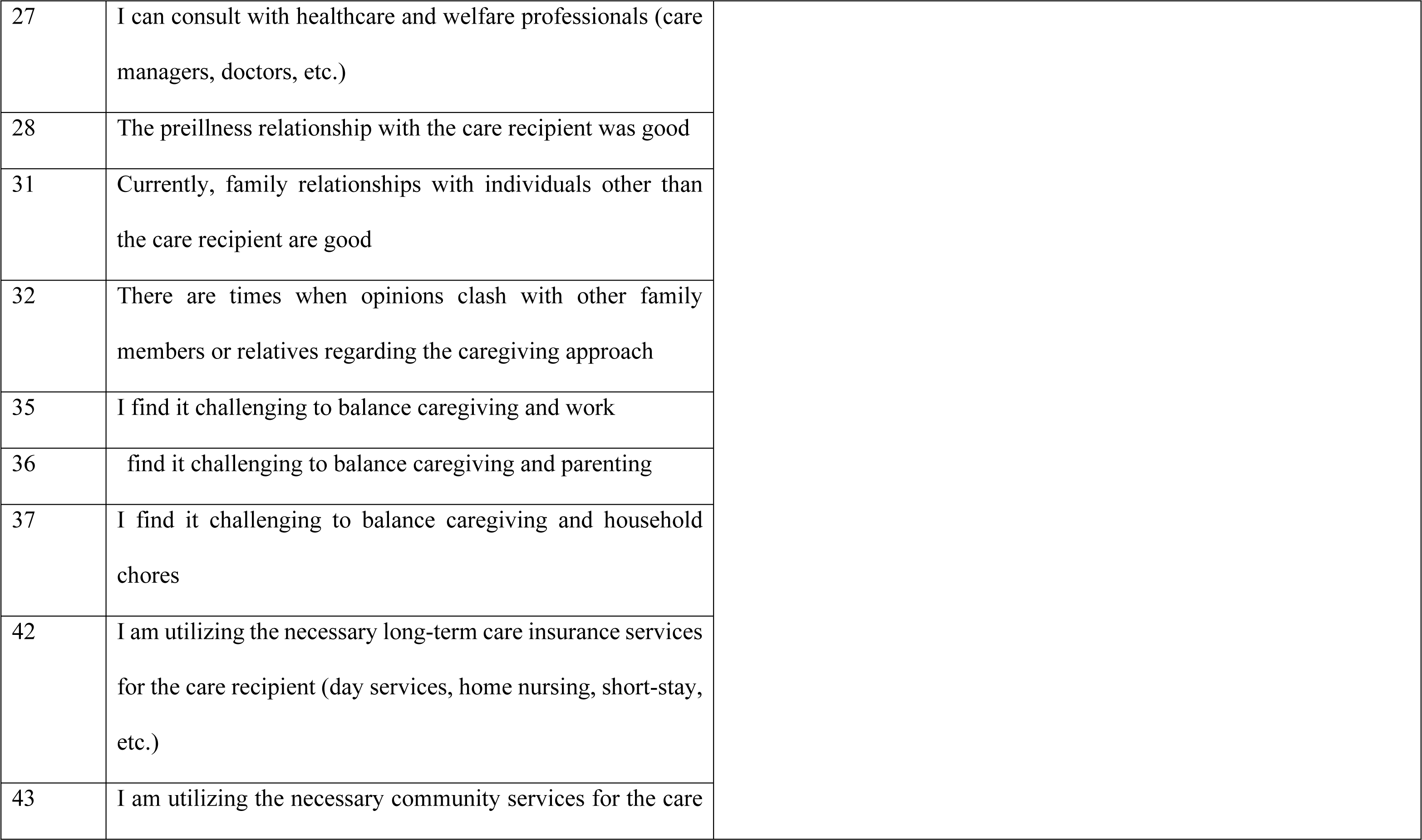
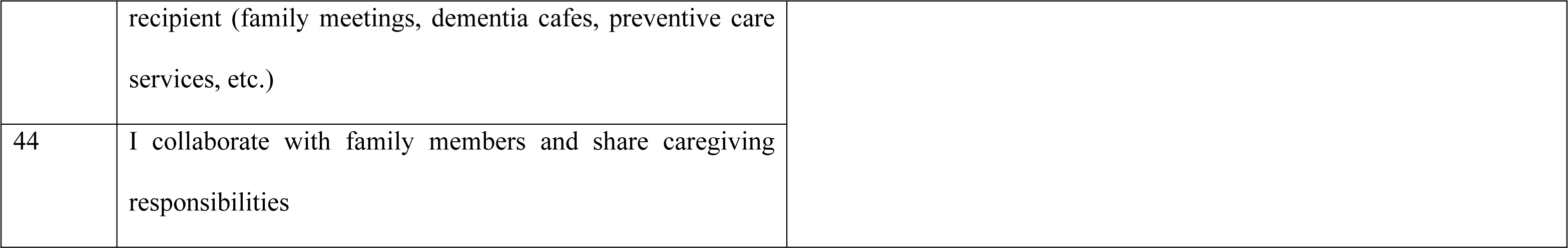
Results of exploratory factor analysis (maximum likelihood method, promax rotation)

|  |  |  | Factor 1 | Factor 2 | Factor 3 | Factor 4 | Factor 5 |
| --- | --- | --- | --- | --- | --- | --- | --- |
|  |  |  | Positive Emotions and Awareness | Existence of Consultation Partners and Family Support | Care Burden and Coping Skills | Dementia Literacy | Involvement and Emotion Control |
| Overall Cronbach's $\alpha$ reliability coefficient for the scale ($\alpha = 0.892$) | | | | | | | |
| Factor 1 Positive Emotions and Awareness ($\alpha = 0.903$) | | | | | | | |
| 20 | Caring for the individual sometimes brings a sense of satisfaction |  | **0.855** | -0.107 | 0.221 | 0.148 | -0.245 |
| 19 | Caring for the individual sometimes uplifts and encourages me |  | **0.774** | −0.078 | 0.195 | 0.043 | −0.098 |
| 24 | Taking care of someone has given meaning to my life |  | **0.771** | 0.027 | −0.030 | −0.041 | 0.039 |
| 23 | I feel happy when I see the person I am caring for enjoying something |  | **0.698** | 0.191 | −0.107 | −0.053 | −0.156 |
| 29 | The current relationship with the care recipient is good |  | **0.665** | 0.113 | −0.068 | −0.064 | 0.217 |
| 25 | I feel that the care recipient is grateful to me |  | **0.659** | 0.129 | −0.164 | 0.001 | −0.005 |
| 18 | I find joy in caregiving at times |  | **0.640** | −0.118 | 0.221 | 0.066 | 0.120 |
| 14 | I can evoke positive responses from the care recipient through verbal communication and caregiving |  | **0.565** | 0.014 | −0.278 | 0.187 | 0.182 |
| 22 | I am providing care not out of a sense of obligation but because I genuinely want to |  | **0.564** | −0.030 | 0.016 | −0.056 | 0.222 |

| **Factor 2 Existence of Consultation Partners and Family Support ($\alpha = 0.802$)** |  |  |  |  |  |  |  |
| --- | --- | --- | --- | --- | --- | --- | --- |
| 39 | I have relatives I can consult with |  | -0.002 | **0.803** | 0.027 | -0.072 | 0.011 |
| 41 | I have neighbors I can consult with |  | 0.043 | **0.684** | -0.03<br>1 | 0.026 | 0.022 |
| 40 | I have friends or acquaintances I can consult with |  | -0.003 | **0.614** | -0.04<br>9 | 0.245 | -0.076 |
| 38 | I have family members I can consult with |  | 0.006 | **0.611** | 0.234 | -0.105 | 0.023 |
| 26 | My family and relatives understand my feelings, including the difficulty of caregiving |  | 0.016 | **0.513** | 0.134 | 0.065 | -0.012 |
| 30 | The family relationships with individuals other than the care recipient were good before the care recipient's illness |  | 0.241 | **0.487** | -0.10<br>6 | -0.159 | 0.020 |
| **Factor 3 Care Burden and Coping Skills ($\alpha = 0.743$)** |  |  |  |  |  |  |  |
| 6 | Feeling stressed due to caregiving taking up time and having no personal time for oneself |  | -0.099 | -0.046 | **0.807** | -0.045 | 0.043 |
| 1 | There are times when I feel restless |  | 0.156 | −0.011 | **0.607** | −0.209 | −0.038 |
| 34 | I can make time for hobbies, travel, and other leisure activities aside from caregiving |  | −0.200 | 0.223 | **0.489** | 0.124 | 0.081 |
| 45 | I consciously make an effort to take regular breaks and engage in activities to avoid caregiver fatigue |  | −0.071 | 0.279 | **0.470** | 0.181 | −0.048 |
| 8 | Sometimes I feel that the caregiving I am doing is not rewarded |  | 0.299 | 0.016 | **0.453** | −0.123 | 0.067 |
| 3 | There are times when I feel down because I cannot interact well with the person receiving care |  | 0.056 | −0.041 | **0.409** | 0.114 | 0.230 |
| **Factor 4 Dementia Literacy ($\alpha = 0.782$)** |  |  |  |  |  |  |  |
| 13 | I have knowledge about the social resources (such as long-term care insurance services) necessary for individuals with dementia |  | −0.002 | −0.027 | 0.106 | **0.832** | 0.029 |
| 12 | I have knowledge about the illness called dementia |  | 0.020 | −0.019 | 0.002 | **0.779** | 0.036 |
| 11 | I can gather information about the knowledge and skills you |  | 0.054 | 0.041 | −0.17 | **0.586** | −0.009 |
|  | feel are lacking in your caregiving |  |  |  | 8 |  |  |
| **Factor 5 Involvement and Emotion Control ($\alpha = 0.783$)** |  |  |  |  |  |  |  |
| 16 | I can respond flexibly to the care recipient's expressions and actions |  | 0.047 | -0.024 | 0.003 | 0.139 | **0.738** |
| 17 | I can provide care to the care recipient in a way that avoids causing discomfort |  | 0.333 | 0.003 | -0.04<br>4 | -0.018 | **0.594** |
| 33 | Even when I feel angry, I can control my emotions |  | 0.041 | 0.024 | 0.237 | -0.082 | **0.576** |
|  | Factor correlation | Factor 1 | 1.000 | 0.232 | 0.381 | 0.224 | 0.581 |
|  |  | Factor 2 |  | 1.000 | 0.141 | 0.275 | 0.222 |
|  |  | Factor 3 |  |  | 1.000 | 0.084 | 0.200 |
|  |  | Factor 4 |  |  |  | 1.000 | 0.399 |
|  |  | Factor 5 |  |  |  |  | 1.000 |
|  |  | Eigenvalues | 7.8 | 2.9 | 2.5 | 2.0 | 1.1 |

|  |  |  |  |  |  |  |
| --- | --- | --- | --- | --- | --- | --- |
|  | Cumulative contribution rate | 27.1 | 35.7 | 43.2 | 48.3 | 51.4 |
| Deleted items |  |  |  |  |  |  |
| 02 | Sometimes I feel irritated when I am around the care recipient |  |  |  |  |  |
| 04 | I feel anxious about taking care of the person for an extended period in the future |  |  |  |  |  |
| 05 | I find myself troubled by the behaviors of the care recipient at times |  |  |  |  |  |
| 07 | Feeling financial burden such as caregiving expenses |  |  |  |  |  |
| 09 | I want to elicit positive responses from the care recipient (such as facial expressions and reduced anxiety) through my caregiving |  |  |  |  |  |
| 10 | It is important to work toward enabling the care recipient to have new roles |  |  |  |  |  |
| 15 | I can understand the care recipient's feelings of anxiety |  |  |  |  |  |
| 21 | There are learning experiences through caregiving |  |  |  |  |  |

|  |  |
| --- | --- |
| 27 | I can consult with healthcare and welfare professionals (care managers, doctors, etc.) |
| 28 | The preillness relationship with the care recipient was good |
| 31 | Currently, family relationships with individuals other than the care recipient are good |
| 32 | There are times when opinions clash with other family members or relatives regarding the caregiving approach |
| 35 | I find it challenging to balance caregiving and work |
| 36 | find it challenging to balance caregiving and parenting |
| 37 | I find it challenging to balance caregiving and household chores |
| 42 | I am utilizing the necessary long-term care insurance services for the care recipient (day services, home nursing, short-stay, etc.) |
| 43 | I am utilizing the necessary community services for the care |
|  | recipient (family meetings, dementia cafes, preventive care services, etc.) |
| 44 | I collaborate with family members and share caregiving responsibilities |

Subsequently, in confirmatory factor analysis, the validity of the five-factor model structure was confirmed with path coefficients of 0.4 or higher from each factor to the observed variables. The model fit indices were CFI = 0.905 and RMSEA = 0.072, meeting the criteria of CFI >0.90 (good), RMSEA of <0.05 (excellent), 0.08 (good), and 0.10 (acceptable).

### Internal consistency

The internal consistency of each subscale was as follows: α = 0.903 for Factor 1, α = 0.802 for Factor 2, α = 0.743 for Factor 3, α = 0.781 for Factor 4, α = 0.783 for Factor 5, and α = 0.892 for the overall scale, all meeting the criteria (Table 4). Structural validity and internal consistency were confirmed, and the CCSD was completed.

## Discussion

### Basic information

The characteristics and caregiving situations of the subjects in this study were compared with those in a nationwide survey conducted in our country [20] to verify their representativeness. The nationwide survey included 3,514 family caregivers, including 789 families after the patient’s death. In the nationwide survey, the average age of caregivers was 62.4 ± 12.2 years; 946 were males (26.9%), and 2,533 were females (72.1%) (with 35 respondents not providing gender information) [20]. In the nationwide survey, among the 2,010 respondents (excluding those who did not respond or were postmortem caregivers), caregiver relationship with the care recipient included spouse (41.6%), biological child (42.5%), son/daughter-in-law (10.1%), sibling (1.1%), and “others” (5%) [20]. In comparison, this study showed a higher average age of caregivers (68.4 ± 9.5 years), with 36.6% of the caregivers being males and 63.3% being females. The majority of caregivers were spouses (61.3%), and biological children accounted for 28.3% (Table 1).

In the nationwide survey, the average age of the care recipients was 81.7 ± 8.7 years [20], which was slightly higher than that of the care recipients targeted in this study. In the nationwide survey, the disease distribution included AD (65.1%), vascular dementia (4.1%), Lewy body dementia (5.0%), frontotemporal dementia (4.2%), mixed-type dementia (1.6%), MCI (1.5%), unknown (10.9%), and others (7.5%) [20]. In contrast, this study had fewer AD cases (Table 1). In the nationwide survey, young-onset dementia was not separately classified, and the AD category in that survey might have included younger-onset cases. Therefore, the distribution of disease types in this study is similar to that of the nationwide survey and within a standard range.

Regarding caregiving situations, the nationwide survey reported that 12.2% of the care recipients were living alone [20], which was comparable to that reported in this study (16.7%). In the nationwide survey, the duration since the diagnosis of dementia was 66 ± 52.8 months, whereas it was 60.0 ± 51.8 months in this study. Regarding the presence of secondary caregivers, in the nationwide survey, among 1,291 respondents, 53.5% had a secondary caregiver, and 36.0% did not [20]. In this study, 42.0% had a secondary caregiver, and 58.0% did not. The higher percentage of “no secondary caregivers” in this study suggests a higher burden on the primary caregiver.

In summary, this study, focusing on elderly caregiving cases where spouses are often caregivers, showed a higher average age of caregivers. Although a direct comparison is challenging because of differences in survey and aggregation methods, an overview of overall attributes and caregiving situations suggests that this study does not significantly deviate from standard data and is comparable to the nationwide survey.

### Structural validity and reliability

Based on the exploratory factor analysis results, considering factors such as Kaiser–Meyer–Olkin, cumulative contribution rate, Cronbach’s α reliability coefficient, and factor loadings of each item, the model with 5 factors and 27 items was determined to be valid. The confirmatory factor analysis results confirmed that this model adequately fits the data obtained from the study participants.

Regarding reliability, Cronbach’s α reliability coefficient for the entire scale was 0.892, indicating internal consistency. Furthermore, all subscales met the criteria for internal consistency.

### Characteristics of each factor

#### Factor 1: Positive emotions and awareness

This factor comprises items related to positive emotions derived from caregiving experiences, positive feelings toward the care recipient, and positive attitudes toward caregiving. The concept of caregiving positivity has been extensively studied since the 1990s, with a focus on caregiving satisfaction [28], meaning in caregiving [29], and self-growth [30]. Furthermore, recent intervention studies have highlighted the importance of caregiving well-being [31] and positive emotions [32,33]. Therefore, this factor was determined to be valid for constituting caregiving competence.

#### Factor 2: Existence of consultation partners and family support

This factor includes items related to the existence of consultation partners, psychological support from family in caregiving situations, and the relationship with family from before the illness. Family functioning [34], the presence of secondary caregivers [35], sharing issues with the community [19], and support from friends [36] have been reported to be associated with caregiving burdens and can be crucial factors in continuing home care. Sharing caregiving situations can strengthen family bonds [37], thus contributing to enhanced caregiving competence.

#### Factor 3: Care burden and coping skills

This factor includes items related to caregiving-related psychological burdens and coping strategies. Previous studies have consistently reported significant psychological stress in dementia caregiving [38–41], and the correlation between the frequency of BPSD expression and psychological stress has been well established [42–44]. Furthermore, numerous studies have emphasized the importance of coping strategies for such stress [45–48], and leisure activities and social interactions with friends have been suggested to play a crucial role in reducing subjective caregiving burden [36]. Based on these findings, this factor was determined to be a valid component in constituting caregiving competence.

#### Factor 4: Dementia literacy

This factor comprises items related to knowledge about dementia, caregiving, and health literacy. In psychological education aimed at reducing BPSD [49], sessions addressing knowledge about the disease [50,51] and caregiving [50,52] are considered crucial. Health literacy is the ability to obtain and process information and services needed to make health-related decisions [53]. Caregivers with high health literacy possess more knowledge about dementia [54] and show a correlation with improved caregiving abilities [55]. The convergence of knowledge about dementia and health literacy contributes to caregiving competence, forming what we term “Dementia Literacy” [56].

#### Factor 5: Involvement and emotion control

This factor includes items related to involvement and emotional control in dementia caregiving situations. These items inquire about subjective perceptions, such as “Do you think you can handle it yourself?” Therefore, they can be interpreted in terms of caregivers’ self-efficacy in dealing with persons with dementia and caregiving tasks. Self-efficacy has an effect on coping strategies [57], and caregivers with stronger self-efficacy can reduce their caregiving burden and enjoy a higher quality of life by taking time for themselves [57,58]. Thus, this factor was considered an important component for controlling caregiving situations and enhancing caregiving competence.

## Conclusion

The structural validity and internal consistency of the CCSD were assessed, and a finalized scale with 5 factors and 27 items that met established criteria was developed. Measuring caregiving competence has the potential to aid in implementing strategies to support caregivers and enable appropriate interventions.

## Data Availability

All relevant data are within the manuscript and its Supporting Information files.

## Author contributions

Ippei Suganuma: Conceptualization, Funding Acquisition, Methodology, Project Administration, Supervision, Visualization, Writing–Original Draft, Writing–Review & Editing

Noriyuki Ogawa: Investigation, Resources

Kenji Kamijou: Investigation, Data Curation

Aki Nakanishi: Methodology

Ippei Kawasaki: Investigation

Keisuke Itotani: Visualization

Shinichi Okada: Formal Analysis, Methodology

## Acknowledgments

We sincerely thank the facilities and family caregivers for their cooperation in collecting the data for this study.

## References

1. World Health Organization. 2023 Mar 15 [cited 2024 Feb 1]. In: Dementia. Available from: https://www.who.int/news-room/fact-sheets/detail/dementia.

2. Statistics Bureau of the Ministry of Internal Affairs and Communications. 2022 Sep 17 [cited 2024 Feb 1]. In: Japan’s Elderly Population from a Statistical Perspective–In Commemoration of ‘Respect for the Aged Day’. Available from: https://www.stat.go.jp/data/topics/topi1380.html.

3. Ministry of Health, Labour and Welfare. White Paper on Aging Society 2017. 2018 [cited 2024 Feb 1]. In: Chapter 1: Situation of Aging (Section 2: Elderly People– Current Status and Trends in their Environment) Available from: https://www8.cao.go.jp/kourei/whitepaper/w-2017/html/gaiyou/s1_2_3.html.

4. Ministry of Health, Labour and Welfare. 2017 [cited 2024 Feb 1]. In: Survey of Household Economy, 2016: IV. Situation of Care. Available from: https://www.mhlw.go.jp/toukei/saikin/hw/k-tyosa/k-tyosa16/dl/05.pdf.

5. Pinquart M, Sörensen S. Differences between caregivers and noncaregivers in psychological health and physical health: a meta-analysis. Psychol Aging. 2003;18: 250–267. doi: 10.1037/0882-7974.18.2.250.

6. Livingston G, Mahoney R, Regan C, Katona C. The caregivers for Alzheimer’s disease Problems Scale (CAPS): development of a new scale within the LASER-AD study. Age Ageing. 2005;34: 287–290. doi: 10.1093/ageing/afi103.

7. Schulz R, O’Brien AT, Bookwala J, Fleissner K. Psychiatric and physical morbidity effects of dementia caregiving: prevalence, correlates, and causes. Gerontologist. 1995;35: 771–791. doi: 10.1093/geront/35.6.771.

8. Markowitz JS, Gutterman EM, Sadik K, Papadopoulos G. Health-related quality of life for caregivers of patients with Alzheimer disease. Alzheimer Dis Assoc Disord. 2003;17: 209–214. doi: 10.1097/00002093-200310000-00003.

9. Kim B, Noh GO, Kim K. Behavioural and psychological symptoms of dementia in patients with Alzheimer’s disease and family caregiver burden: a path analysis. BMC Geriatr. 2021;21: 160.doi: 10.1186/s12877-021-02109-w.

10. Alzheimer’s Association. 2023 Alzheimer’s disease facts and figures. Alzheimers Dem. 2023;19: 1598–1695. doi: 10.1002/alz.13016.

11. Brodaty H, Hadzi-Pavlovic D. Psychosocial effects on carers of living with persons with dementia. Aust N Z J Psychiatry. 1990;24: 351–361. doi: 10.3109/00048679009077702.

12. Gallicchio L, Siddiqi N, Langenberg P, Baumgarten M. Gender differences in burden and depression among informal caregivers of demented elders in the community. Int J Geriatr Psychiatry. 2002;17: 154–163. doi: 10.1002/gps.538.

13. Schulz R, Williamson GM. A 2-year longitudinal study of depression among Alzheimer’s caregivers. Psychol Aging. 1991;6: 569–578. doi: 10.1037//0882-7974.6.4.569.

14. Ministry of Health, Labour and Welfare. Manual for Family Caregiver Support by Municipal Comprehensive Support Centers: Support for Caregivers’ Life Journey. 2017 [cited 2024 Feb 1]. In: The Urgent Need for Advancing Support Policies and Programs for Family Caregivers with a Fresh Perspective. Available from: chrome-extension://efaidnbmnnnibpcajpcglclefindmkaj/https://www.mhlw.go.jp/content/12300000/000307003.pdf.

15. Folkman S, Moskowitz JT. Stress, positive emotion, and coping. Curr Dir Psychol Sci. 2000;9: 115–118. doi: 10.1111/1467-8721.00073.

16. Fredrickson BL. What good are positive emotions? Rev Gen Psychol. 1998;2: 300–319. doi: 10.1037/1089-2680.2.3.300.

17. Tugade MM, Fredrickson BL, Barrett LF. Psychological resilience and positive emotional granularity: examining the benefits of positive emotions on coping and health. J pers. 2004;72: 1161–1190. doi: 10.1111/j.1467-6494.2004.00294.x.

18. Wu X. Main care-givers empowerment measurement (MCEM). J Jpn Acad Nurs Sci, Chinese version. 2008;28: 3–13 (in Japnanese).

19. Sakanashi S and Fujita K. Development of the empowerment scale for family caregivers of community-dwelling people with dementia in Japan. Jpn J Nurs Sci. 2020;17: e12311. doi: 10.1111/jjns.12311.

20. Suzuki M Survey on the Thoughts of Families of People with Dementia and the Support They Receive: A Real-World Investigation. “Subsidy for the Promotion of Elderly Health Project in the Fiscal Year 2021: Senior Health Promotion and Health Enhancement Grant –”. In: Survey Results on Support for Families of per with Dementia, Hara N, Inomata S, Shibuya M, Okura Y. Kyoto: Alzheimer’s Association Japan; 2022. pp. 26–67.

21. Liew TM, Tai BC, Yap P, Koh GC. Contrasting the risk factors of grief and burden in caregivers of persons with dementia: multivariate analysis. Int J Geriatr Psychiatry. 2019;34: 258–264. doi: 10.1002/gps.5014.

22. Rosdinom R, Zarina MZ, Zanariah MS, Marhani M and Suzaily W. Behavioural and psychological symptoms of dementia, cognitive impairment and caregiver burden in patients with dementia. Prev Med. 2013;57: S67–S69. doi: 10.1016/j.ypmed.2012.12.025.

23. Watson B, Tatangelo G, McCabe M. Depression and anxiety among partner and offspring carers of people with dementia: a systematic review. Gerontologist. 2019;59: e597–e610. doi: 10.1093/geront/gny049.

24. Shima S, Shikano T, Kitamura T, Asai M. New self-rating scales for depression New depressive self-rating scale. Clin Psychiatry. 1985;27: 717–723. (in Japanese).

25. Furuta K, Tanaka T, Ogisawa F, Matsui H, Omori Y, Awata S. Study on equivalence of the revised short-form version of Dementia Behavior Disturbance Scale. Jpn J. 2022;59: 384–387. (in Japanese).

26. Kline RB. Principles and practice of structural equation modeling, 2nd ed. New York: Guilford Press; 2005.

27. Clark LA, Watson D. Constructing validity: basic issues in objective scale development. Psychol Assess. 1995;7: 309–319. doi: 10.1037/1040-3590.7.3.309.

28. Kramer BJ. Gain in the caregiving experience: where are we? What next? Gerontologist. 1997;37: 218–232. doi: 10.1093/geront/37.2.218.

29. Farran CJ, Keane-Hagerty E, Salloway S, Kupferer S, Wilken CS. Finding meaning: an alternative paradigm for Alzheimer’s disease family caregivers. Gerontologist. 1991;31: 483–489. doi: 10.1093/geront/31.4.483.

30. Skaff MM, Pearlin LI. Caregiving: role engulfment and the loss of self. Gerontologist. 1992;32: 656–664. doi: 10.1093/geront/32.5.656.

31. Gonçalves AC, Demain S, Samuel D, Marques A. Physical activity for people living with dementia: carer outcomes and side effects from the perspectives of professionals and family carers. Aging Clin Exp Res. 2021;33: 1267–1274. doi: 10.1007/s40520-020-01636-7.

32. Yoon HK, Kim GS. An empowerment program for family caregivers of people with dementia. Public Health Nurs. 2020;37: 222–233. doi: 10.1111/phn.12690.

33. Suganuma I, Segawa D, Kamijou K, Okada S. Study of support for enhancing empowerment of primary family caregiver who cares for elderly persons with dementia: a pilot study of a psychoeducational intervention. The Jpn J Occup Ther. 2021;55: 1529–1535 (in Japanese).

34. Yu H, Wang X, He R, Liang R, Zhou L. Measuring the caregiver burden of caring for community-residing people with Alzheimer’s disease. PLoS One. 2015;10: e0132168. doi: 10.1371/journal.pone.0132168. eCollection.

35. Kim B, Kim JI, Na HR, Lee KS, Chae KH, Kim S. Factors influencing caregiver burden by dementia severity based on an online database from Seoul dementia management project in Korea. BMC Geriatr. 2021;21: 649. doi: 10.1186/s12877-021-02613-z.

36. Steinsheim G, Malmedal W, Follestad T, Olsen B, Saga S. Factors associated with subjective burden among informal caregivers of home-dwelling people with dementia: a cross-sectional study. BMC Geriatr. 2023;23: 644. doi: 10.1186/s12877-023-04358-3.

37. Uchiyama S, Tsukada N, Sakuraba T. The family support by family psycho education based on EBP in community mental disabled facilities. Kanto Gakuin Univ J Nurs. 2015;2: 11-20.

38. Schoenmakers B, Buntinx F, Delepeleire J. Factors determining the impact of care-giving on caregivers of elderly patients with dementia. A systematic literature review. Maturitas. 2010;66: 191–200. doi: 10.1016/j.maturitas.2010.02.009.

39. Sheehan OC, Haley WE, Howard VJ, Huang J, Rhodes JD, Roth DL. Stress, burden, and well-being in dementia and nondementia caregivers: insights from the caregiving transitions study. Gerontologist. 2021;61: 670–679. doi: 10.1093/geront/gnaa108.

40. American Association of Retired Persons. Caring for people with dementia: Caregivers’ experiences. 2018 Nov [cited 2024 Feb 1]. In: Caregivers Cite Emotions and the Demands of Care as the Biggest Challenges of Caring for Someone with Dementia. Available from: https://www.aarp.org/content/dam/aarp/research/surveys_statistics/ltc/2018/caring-people-dementia-survey.doi.10.26419-2Fres.00262.001.pdf.

41. Goto Y, Morita K, Suematsu M, Imaizumi T and Suzuki Y. Caregiver burdens, health risks, coping and interventions among caregivers of dementia patients: a review of the literature. Intern Med. 2023;62: 3277–3282. doi: 10.2169/internalmedicine.0911-22.

42. de Vugt ME, Nicolson NA, Aalten P, Lousberg R, Jolle J, Verhey FRJ. Behavioral problems in dementia patients and salivary cortisol patterns in caregivers. J Neuropsychiatry Clin Neurosci. 2005;17: 201–207. doi: 10.1176/jnp.17.2.201.

43. Savla J, Roberto KA, Blieszner R, Cox M, Gwazdauskas F. Effects of daily stressors on the psychological and biological well-being of spouses of persons with mild cognitive impairment. J Gerontol B Psychol Sci Soc Sci. 2011;66: 653–664. doi: 10.1093/geronb/gbr041.

44. Savla J, Granger DA, Roberto KA, Davey A, Blieszner R, Gwazdauskas F. Cortisol, alpha amylase, and daily stressors in spouses of persons with mild cognitive impairment. Psychol Aging. 2013;28: 666-679. doi: 10.1037/a0032654.

45. Almberg B, Grafström M, Winblad B. Major strain and coping strategies as reported by family members who care for aged demented relatives. J Adv Nurs. 1997;26: 683–691. doi: 10.1046/j.1365-2648.1997.00392.x.

46. Gilhooly KJ, Gilhooly ML, Sullivan MP, McIntyre A, Wilson L, Harding E, et al. A meta-review of stress, coping and interventions in dementia and dementia caregiving. BMC Geriatr. 2016;16: 106. doi: 10.1186/s12877-016-0280-8.

47. Monteiro AMF, Santos RL, Kimura N, Baptista MAT, Dourado MCN. Coping strategies among caregivers of people with Alzheimer disease: a systematic review. Trends Psychiatry Psychother. 2018;40: 258–268. doi: 10.1590/2237-6089-2017-0065.

48. Cooper C, Katona C, Orrell M, Livingston G. Coping strategies, anxiety and depression in caregivers of people with Alzheimer’s disease. Int J Geriatr Psychiatry. 2008;23: 929–936. doi: 10.1002/gps.2007.

49. Livingston G, Johnston K, Katona C, Paton J, Lyketsos CG and Old Age Task Force of the World Federation of Biological Psychiatry. Systematic review of psychological approaches to the management of neuropsychiatric symptoms of dementia. Am J Psychiatry. 2005;162: 1996–2021. doi: 10.1176/appi.ajp.162.11.1996.

50. Llanque SM, Enriquez M, Cheng AL 3, Doty L, Brotto MA, Kelly PJ, et al. The family series workshop: a community-based psychoeducational intervention. Am J Alzheimers Dis Other Demen. 2015;30: 573–583.

51. Ponce CC, Ordonez TN, Lima-Silva TB, Dos Santos GD, Viola LF, Nunes PV, et al. Effects of a psychoeducational intervention in family caregivers of people with Alzheimer’s disease. Dement Neuropsychol. 2011;5: 226–237. doi: 10.1590/S1980-57642011DN05030011.

52. Livingston G, Barber J, Rapaport P, Knapp M, Griffin M, King D, et al. Clinical effectiveness of a manual based coping strategy programme (START, STrAtegies for RelaTives) in promoting the mental health of carers of family members with dementia: pragmatic randomised controlled trial. BMJ. 2013;347: f6276. doi: 10.1136/bmj.f6276.

53. Nielsen-Bohlman L, Panzer AM, Kindig DA, eds. Institute of Medicine (US) committee on health literacy. Washington (DC): National Academies Press. US; 2004.

54. Crawley S, Moore K, Vickerstaff V, Fisher E, Cooper Cl, Sampson EL. How do factors of sociodemographic, health literacy and dementia experience influence carers’ knowledge of dementia? Dementia (London). 2022;21: 1270–1288. doi: 10.1177/14713012221074219.

55. Li Y, Hu L, Mao X, Shen Y, Xue H, Hou P, et al. Health literacy, social support, and care ability for caregivers of dementia patients: structural equation modeling. Geriatr Nurs. 2020;41: 600–607. doi: 10.1016/j.gerinurse.2020.03.014.

56. Lo RY. Uncertainty and health literacy in dementia care. Ci Ji Yi Xue Za Zhi. 2020;32: 14–18. doi: 10.4103/tcmj.tcmj_116_19.

57. Gonyea JG, O’Connor M, Carruth A, Boyle PA. Subjective appraisal of Alzheimer’s disease caregiving: the role of self-efficacy and depressive symptoms in the experience of burden. Am J Alzheimers Dis Other Demen. 2005;20: 273–280. doi: 10.1177/153331750502000505.

58. Coen RF, O’Boyle CA, Coakley D, Lawlor BA. Individual quality of life factors distinguishing low-burden and high-burden caregivers of dementia patients. Dement Geriatr Cogn Disord. 2002;13: 164–170. doi: 10.1159/000048648.

